# CNS-selective plasma p-tau217 accurately captures Alzheimer’s disease pathology and progression

**DOI:** 10.64898/2026.05.18.26353208

**Authors:** Xuemei Zeng, Marissa F. Farinas, Michel N. Nafash, Denis S. Smirnov, Ruyu Shi, Brian J. Lopresti, Cristy Matan, Dana L. Tudorascu, Sarah B. Berman, Robert A. Sweet, C. Elizabeth Shaaban, Beth E. Snitz, Julia K. Kofler, Neelesh K. Nadkarni, Howard J. Aizenstein, Alzheimer’s Disease Neuroimaging Initiative, Milos D. Ikonomovic, Tharick A. Pascoal, Victor L. Villemagne, M. Ilyas Kamboh, Henrik Zetterberg, Ann D. Cohen, Oscar L. Lopez, Thomas K. Karikari

## Abstract

Blood-based biomarkers are transforming Alzheimer’s disease (AD) detection, with plasma tau phosphorylated at threonine 217 (p-tau217) being a leading candidate. However, circulating p-tau217 originates from both central nervous system (CNS) and non-CNS sources, limiting specificity. Here, we benchmark the performance of next-generation p-tau217 biomarkers that are selective for p-tau217 from predominantly CNS origin versus the conventional non-selective p-tau217. Across neuropathologically-defined, Aβ- and tau-neuroimaging–characterized, and memory clinic cohorts, plasma brain-derived-p-tau217 (BD-p-tau217)–either alone or in ratio with brain-derived tau expressed as a percentage (BD-p-tau217%)–identified AD pathology and clinical AD with larger effect sizes, higher discriminative accuracies, and improved sensitivity and specificity, outperforming conventional alternatives. Furthermore, BD-p-tau217% and BD-p-tau217 demonstrated more robust prediction of future clinical progression up to two decades later. Importantly, diagnostic performance remained high in older adults with diabetes and cardiovascular disease, populations in which standard p-tau217 showed reduced specificity. The findings were independently replicated in a large multicenter cohort, demonstrating the robustness and generalizability of the observed improvements across clinical, imaging, and longitudinal outcomes. These results establish plasma BD-p-tau217% and BD-p-tau217 as biologically grounded and clinically robust biomarkers that advance early detection, diagnosis and prognosis of AD.

**One Sentence Summary:** Plasma brain-derived p-tau217 and its ratio over brain-derived tau improve the specificity of the leading blood-based Alzheimer’s disease biomarker p-tau217.

## INTRODUCTION

Alzheimer’s disease (AD) remains the most common cause of dementia worldwide, affecting 55 million individuals globally and costing over 1.3 trillion US dollars (*1*). AD pathology – with the hallmark features of amyloid-beta (Aβ) plaques and tau neurofibrillary tangles (NFTs) – starts to manifest years before clinical symptoms appear, and many more years before dementia onset (*2–4*). This clinical-biological disconnect necessitates the development and application of high-performance biomarkers to enable early and accurate detection, prediction and monitoring, as well as to support therapeutic and observational trials (*5–8*). While molecular imaging and cerebrospinal fluid (CSF) biomarkers fulfill these criteria, their limited scalability, high costs and invasiveness are notable limitations against widespread implementation in specialist clinical settings as well as in primary care and population-based screenings (*9–12*). In recent years, well-validated blood biomarkers have become available as viable alternatives with added advantages of scalability, accessibility and cost effectiveness (*11–13*). Among these, plasma tau phosphorylated at threonine 217 (p-tau217) has shown the highest diagnostic accuracies, and the strongest and most consistent correspondence with brain Aβ and tau NFT pathology (*14–17*). Consequently, clinical practice guidelines recommend p-tau217 testing as an integral part of the clinical diagnostic and prognostic workup for AD (*18, 19*). Furthermore, commercial plasma p-tau217 tests with regulatory approval or Laboratory Developed Test status are now available, facilitating clinical uptake (*20–24*).

Despite the high performance, plasma p-tau217 testing has its own challenges (*25*). For example, plasma p-tau217 diagnostic accuracies are substantially decreased in individuals with common comorbidities of aging such as hypertension, myocardial infarction, diabetes, chronic kidney disease (CKD), hypercholesterolemia, and obesity (*26–28*). Moreover, although it is highly specific for AD in the context of neurodegenerative diseases, plasma p-tau217 is also elevated in non-AD conditions such as amyotrophic lateral sclerosis (ALS), HIV-associated neurocognitive disorders, Niemann Pick disease type C, systemic amyloidosis and CKD (*29–33*). These findings can be partly explained by the presence of p-tau217 signal not only in central nervous system (CNS) tissue (*34, 35*) but also in peripheral tissues such as muscle biopsies of ALS patients and unaffected controls (*29*) and in blood samples from individuals diagnosed with polyneuropathy due to systemic amyloidosis (*33*). The reduced biological specificity and diagnostic precision caused by these mixed-origin measurements raises concern that conventional plasma p-tau217 may not exclusively capture CNS-derived neuropathology.

A central challenge, therefore, is to distinguish CNS-derived pathological tau species from peripheral background signal. Tau, encoded by the *MAPT* gene, is expressed as multiple isoforms that can be broadly divided into low-molecular-weight (LMW) and high-molecular-weight (HMW, or big tau) forms based on inclusion of exon 4a (*36, 37*). LMW tau isoforms constitute the predominant tau species in the CNS (*38*), whereas big tau is primarily expressed in the peripheral nervous system (PNS) and peripheral organs such as the heart and liver (*39–42*). We therefore hypothesized that selectively quantifying circulating LMW tau phosphorylated at threonine 217, a brain-enriched species, would enhance biological specificity and clinical performance.

To test this hypothesis, we employed the Nucleic acid Linked Immuno-Sandwich Assay (NULISA™) proximity ligation proteomic platform (*43*) to quantify CNS-relevant plasma biomarkers for optimal AD diagnosis (see Data S1 for biomarker list). Using this platform, we evaluated the clinical performance of plasma brain-derived-p-tau217 (BD-p-tau217) and BD-p-tau217% vs. other biomarkers. While BD-p-tau217 is a single immunoassay, BD-p-tau217% takes a ratio of BD-p-tau217 to the total amount of phosphorylation-independent CNS-derived tau protein (i.e., BD-tau) in plasma expressed as a percentage, leveraging our innovation of plasma BD-tau as a biomarker of CNS-selective tau protein in circulating blood (*44, 45*) (Figure S1). We demonstrate that BD-p-tau217% and BD-p-tau217 outperform conventional plasma p-tau217, p-tau217/Aβ42 (an Food and Drugs Administration [FDA] approved plasma AD biomarker), and p-tau217% (non-CNS-selective immunoassay alternative to BD-p-tau217%, calculated as a ratio of p-tau217/total tau) originally described by Barthélemy et al (*46*), across multiple diagnostic contexts, including in individuals with common age-related comorbidities, and provides superior prediction of future clinical progression.

## RESULTS

### Cohort characteristics

We selected three cohorts (clinical [n=1386], neuropathology [n=323], and neuroimaging [n=242]) from the University of Pittsburgh Alzheimer’s Disease Research Center (Pitt ADRC) academic memory clinic. The cohorts included participants recruited between 1993 and 2023 and monitored clinically for up to two decades. Figure S2 summarizes the participant selection criteria, relationship between cohorts, and the derivation of sub cohorts employed in longitudinal analyses. The clinical cohort comprised 53.0% females, 91.5% non-Hispanic Whites, and 46.7% *APOE4* carriers; these characteristics were similar in the neuropathology and neuroimaging cohorts. The median (interquartile range [IQR]) age at blood draw was 71.4 (12.8), 68.5 (12.8) and 74.5 (14.6) years for the clinical, neuroimaging and neuropathology cohorts, respectively.

The neuropathology cohort included individuals with matched antemortem clinical and biomarker assessments in addition to postmortem autopsy diagnosis. Median (IQR) blood-to-death interval was 5.9 (5.4) years, and there were 17.3%, 14.6% and 68.1% participants with low, intermediate and severe Alzheimer’s disease neuropathological change (ADNC), respectively. The neuroimaging cohort included 242 participants who underwent positron emission tomography (PET) imaging for Aβ plaques (Aβ-PET; 53.1% positive) and/or NFTs (tau-PET; 54.1% positive). The clinical cohort, focusing on clinical diagnoses alone, included 29.7% clinically unimpaired (CU), 20.7% mild cognitive impairment (MCI), 34.6% AD dementia (probable/possible AD), and 14.9% non-AD dementia. Table 1 summarizes participant characteristics for cohorts while Tables S1-3 show cohort-specific characteristics.

**Table 1.** Baseline characteristics of the Pitt-ADRC study participants.

| **Characteristics** | **Neuropathology cohort** | **Neuroimaging cohort** | **Clinical cohort** |
|---|---|---|---|
| Number of participants, n | 323 | 242 | 1386 |
| Age (blood; years), median (IQR) | 74.5 (14.6) | 68.5 (12.8) | 71.4 (12.8) |
| Blood-to-death (years), median (IQR) | 5.3 (3.6) | NA | NA |
| Female, n (%) | 158 (48.9%) | 104 (43.0%) | 735 (53.0%) |
| NHW, n (%) | 316 (97.8%) | 232 (95.9%) | 1268 (91.5%) |
| Education (years), median (IQR) | 15 (5) | 16 (4) | 16 (6) |
| *APOE4* carriers, n (%) [missing n] | 173 (53.7%) [1] | 95 (41.0%) [10] | 639 (46.7%) [16] |
| MMSE (baseline), median (IQR) | 21 (8) | 27 (6) | 26 (7) |
| Clinical diagnosis, n (%) |  |  |  |
| CU | 12 (3.8%) | 64 (27.6%) | 412 (29.7%) |
| MCI | 21 (6.7%) | 70 (30.2%) | 287 (20.7%) |
| AD dementia | 238 (75.6%) | 72 (31.0%) | 480 (34.6%) |
| Non-AD dementia | 44 (14.0%) | 26 (11.2%) | 207 (14.9%) |
| Missing | 8 | 10 | 0 |
| Aβ-PET (Neg/Pos/Missing), n | Not applicable | 113/128/1 | Not applicable |
| Tau-PET (Neg/Pos/Missing), n | Not applicable | 39/46/157 | Not applicable |
| ADNC (Low/Int/High), n | 56/47/220 | Not applicable | Not applicable |
| NFT Braak (Low/PART/Mid/High), n | 34/11/47/220 | Not applicable | Not applicable |
| CERAD rating (Low/Mod/Freq), n | 48/23/252 | Not applicable | Not applicable |

**Abbreviation:** IQR, inter-quartile range; NHW, Non-Hispanic White; AD, Alzheimer’s disease; CU, cognitively unimpaired; MCI, mild cognitive impairment; PET, positron emission tomography; MMSE, Mini-Mental State Examination; ADNC, Alzheimer’s disease neuropathologic change; NFT, neurofibrillary tangle. ADNC was categorized as low, intermediate, or high; NFT Braak stage as low, PART (primary age□related tauopathy), middle, or high; and CERAD neuritic plaque density rating as low, moderate, or frequent.

Additionally, we included 1428 participants (529 CU, 676 MCI, 223 dementia) from the Alzheimer’s Disease Neuroimaging Initiative (ADNI; phase 1 to 4) as an external validation cohort. The median age was 72.8 (10.3) years, consisting of 47.3% female, 95.3% non-Hispanic White, and 45.4% *APOE4* carriers. Aβ-PET data were available for 999 participants (49.5% positive), and tau-PET data for 360 participants (20.6% positive). Participant characteristics for the overall cohort and stratification by Aβ-PET, tau-PET and clinical diagnosis, are summarized in Tables S4-S7.

### Discriminative accuracy for ADNC in the Pitt-ADRC neuropathology cohort

Plasma BD-p-tau217% was the topmost elevated biomarker which best distinguished the low, intermediate and severe ADNC cases, followed by BD-p-tau217 (Fig. 1). For the primary analyses discriminating low ADNC (i.e., minimal or no AD related neuropathologic change) from severe ADNC (definite AD diagnosis), BD-p-tau217% had the largest age- and sex-adjusted Cohen’s *d* effect size of 2.53, relative to 2.12, 1.87, 1.77 and 1.86 for BD-p-tau217, p-tau217%, p-tau217 and p-tau217/Aβ42, respectively (Fig. 1A). BD-p-tau217% and BD-p-tau217 achieved equivalent AUCs of 0.93 each, both being significantly higher than the AUC of 0.90 (95%CI=0.83–0.93) for p-tau217 (p=0.003) (Fig. 1A and Data S2-3). Although p-tau217% showed a numerically higher AUC than p-tau217, the difference was not significant. BD-p-tau217% correctly classified 93% of individuals, compared with 85%-90% for the other biomarkers.

**Figure 1.**
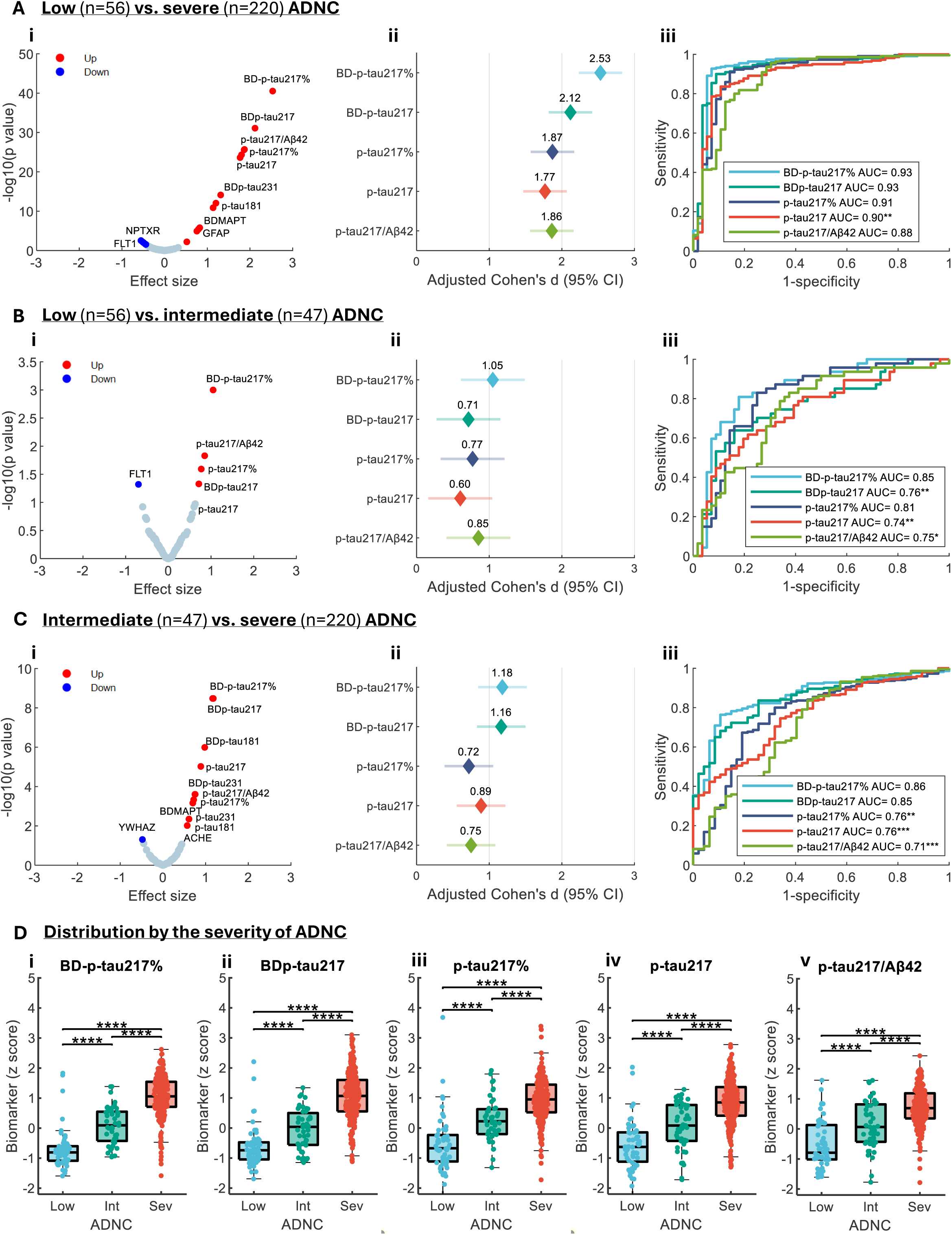
Association of plasma biomarker levels with autopsy-defined ADNC in the Pitt-ADRC neuropathology cohort. Panels A-C compare biomarker levels between low vs. severe ADNC, low vs. intermediate ADNC, and intermediate vs. severe ADNC groups, respectively. Volcano plots display age and sex adjusted Cohen’s d effect sizes on the x axis and -log (p values) on the y axis. Forest plots depict the point estimates and 95% confidence intervals of Cohen’s d. Biomarker levels were z-normalized. AUC plots were based on logistic regression classification performance using biomarker level alone. Asterisks denote DeLong test comparisons with BD-p tau217% as the reference biomarker, with *, **, and *** indicating p < 0.05, 0.01, and 0.001, respectively. Panels D show boxplot distributions of biomarker levels across ADNC categories; the center line indicates the median, box edges represent the 25th and 75th percentiles, and whiskers extend to the most extreme non outlier values. Asterisks denote significance levels for pairwise rank sum tests after Benjamini–Hochberg correction, with *, **, ***, and **** for p < 0.05, 0.01, 0.001, and 0.0001, respectively.

In secondary analyses differentiating low from intermediate ADNC, BD-p-tau217% was again the best-performing biomarker with the highest effect size (1.05, vs. 0.60-0.85 for all others; Fig. 1B). BD-p-tau217% achieved an AUC of 0.85 (95%CI=0.75–0.91; 82% correctly classified), yielding a higher AUC than p-tau217% (0.81[0.70-0.88]; 77% correctly classified), and significantly outperforming each of BD-p-tau217 (0.76[95%CI=0.65–0.85; 75% correctly classified]; p=0.002), p-tau217/Aβ42 (0.75[95%CI=0.62–0.82; 72% correctly classified]; p=0.021) and p-tau217 (0.74[95%CI=0.63–0.83; 71% correctly classified]; p=0.005) (Fig. 1B and Data S2-3).

Similar results were recorded when comparing intermediate vs. severe ADNC (Fig. 1C and Data S2-3). Plasma BD p tau217% and BD p tau217 achieved equivalent AUCs of 0.86 (95%CI=0.79–0.90) and 0.85 (95%CI=0.79–90) respectively, both significantly outperforming p tau217%, p tau217, and p tau217/Aβ42, whose AUCs ranged from 0.71 to 0.76 (p<0.01). These results were corroborated by boxplots showing the least group-level overlaps between ADNC categories for BD-p-tau217% and BD-p-tau217 vs. the other analytes (Fig. 1D).

Consistent results were recorded when the neuropathology cohort was assessed according to brain NFT Braak stage (Fig. 2) and neuritic plaque density (Fig. S3). Plasma BD-p-tau217% was the topmost hit and had the largest effect sizes and highest AUCs across all pathology comparisons, except for contrasting between moderate and frequent neuritic plaque density where all biomarkers performed equivalently (Fig. S3), and between primary age related tauopathy (PART) and mid Braak stages where BD-p-tau217% outperformed BD-p-tau217 and p-tau217 but not the ratio-based biomarkers i.e., p-tau217/Aβ42 and p-tau217% (Fig. 2). BD p tau217% demonstrated significantly higher AUCs than p tau217 for distinguishing low from frequent neuritic plaque density, as well as for all comparisons across NFT Braak stages. BD p tau217% also outperformed BD p tau217 in distinguishing PART from mid Braak.

**Fig. 2.**
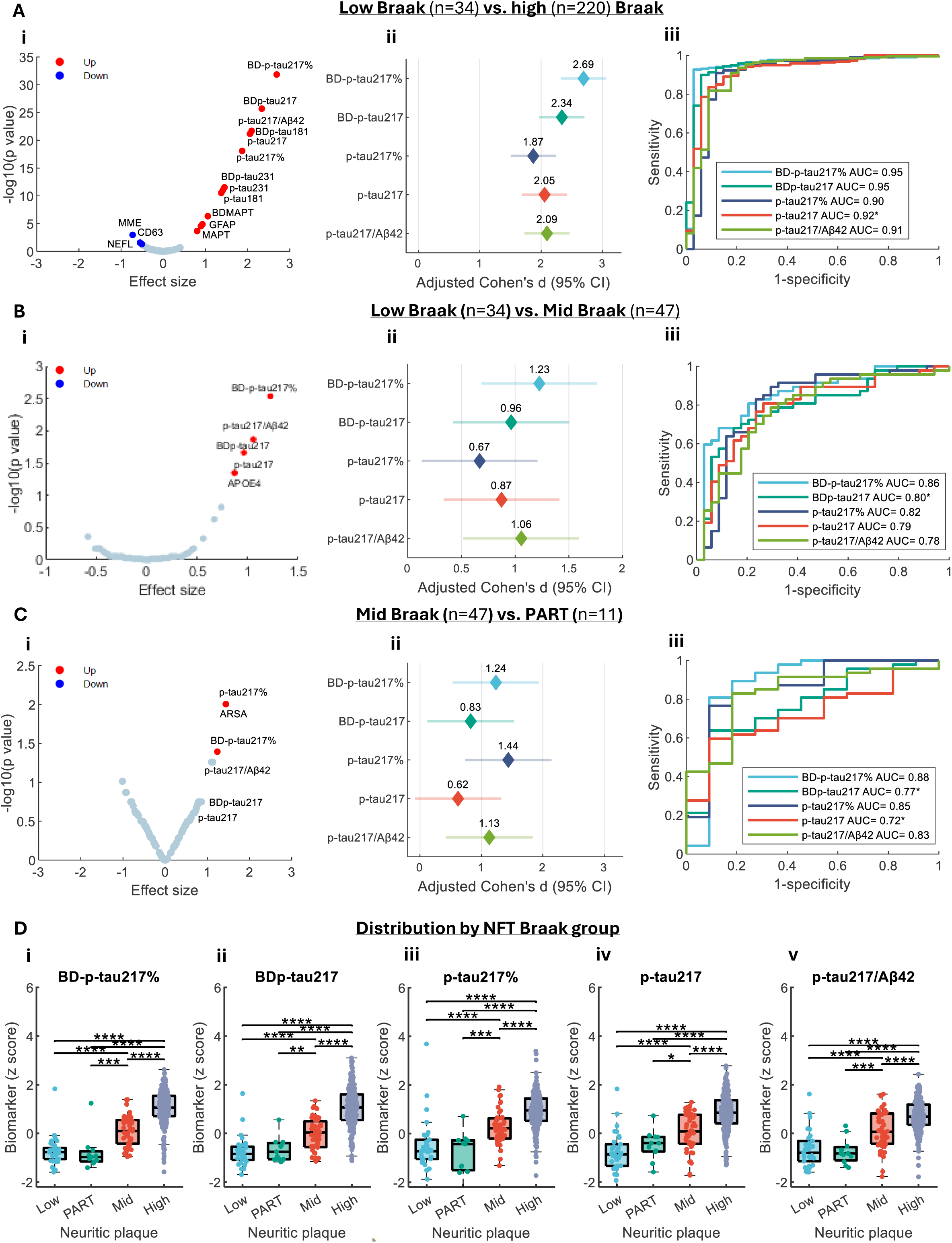
Association of biomarker levels with NFT Braak stage in the Pitt-ADRC neuropathology cohort. Panels A–C compare biomarker levels between low-Braak vs. high-Braak, low-Braak vs. mid-Braak, and mid-Braak vs. PART (primary age-related tauopathy), respectively. Volcano plots display age and sex adjusted Cohen’s d effect sizes on the x axis and -log (p values) on the y axis. Forest plots depict the point estimates and 95% confidence intervals of Cohen’s d. Biomarker levels were z-normalized. AUC plots were based on logistic regression classification performance using biomarker level alone. Asterisks denote DeLong test comparisons with BD-p tau217% as the reference biomarker, with *, **, and *** indicating p < 0.05, 0.01, and 0.001, respectively. Panel D show boxplot distributions of biomarker levels across NFT Braak stages; the center line indicates the median, box edges represent the 25th and 75th percentiles, and whiskers extend to the most extreme non-outlier values. *, **, ***, and **** are for p < 0.05, 0.01, 0.001, and 0.0001, respectively. P values were FDR-adjusted using Benjamini-Hochberg procedure.

### Relation with *in vivo* Aβ and tau PET status in the Pitt-ADRC neuroimaging cohort

Plasma BD-p-tau217% levels best discriminated abnormal vs. normal Aβ-PET (A+ vs. A-) groups, with the greatest effect size (2.18 vs. 1.65 for p-tau217, 1.53 for p-tau217% and 1.34 for p-tau217/Aβ42) and an AUC of 0.94(95%CI=0.90–0.96; 90% correctly classified). This performance was significantly stronger (p<0.001) than the AUCs of 0.87(95%CI=0.81–0.91; 80% correctly classified), 0.87(95%CI=0.82–0.92; 83% correctly classified), and 0.82(95%CI=0.76–0.87; 76% correctly classified) for p-tau217, p-tau217%, and p-tau217/Aβ42, respectively, but not BD-p-tau217 (0.93; 95%CI=0.89-0.95; Fig. 3A-B and Data S2-3).

**Figure 3.**
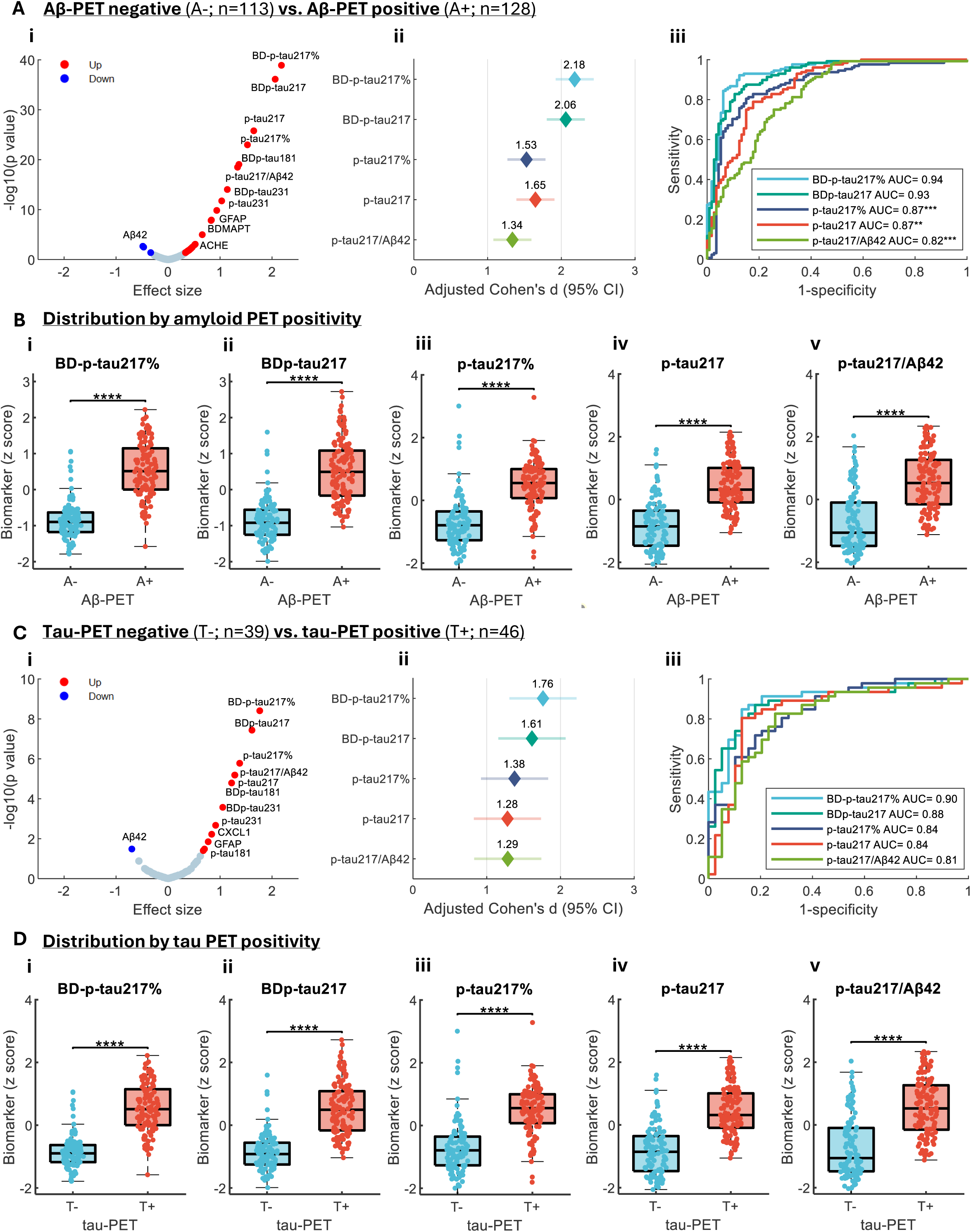
Association of plasma biomarker levels with PET-defined AD pathology in the Pitt-ADRC neuroimaging cohort. Panels A-C show comparison results for Aβ PET negative vs. Aβ PET positive (A) and tau PET negative vs. tau PET positive (C). Volcano plots display age and sex adjusted Cohen’s d effect sizes on the x axis and −log (p values) on the y axis. Red dots indicate biomarkers up regulated, and blue dots indicate biomarkers down regulated, in the PET positive groups. P values were FDR adjusted using the Benjamini–Hochberg procedure. Forest plots show point estimates and 95% confidence intervals of Cohen’s d. Biomarker levels were z normalized. AUC plots illustrate logistic regression classification performance using biomarker level alone. Asterisks denote DeLong test comparisons using BD-p tau217% as the reference biomarker, with *, **, and *** indicating p < 0.05, 0.01, and 0.001, respectively. Panels B and D display box plot distributions of biomarker levels by Aβ PET status (B) and tau PET status (D). The center line indicates the median, box edges correspond to the 25th and 75th percentiles, and whiskers extend to the most extreme non outlier values. All pairwise rank sum tests yielded Benjamini–Hochberg–corrected p < 0.0001, denoted by ****.

Regarding separating tau PET-positive (T+) from tau PET-negative (T-) individuals, BD-p-tau217% had the greatest effect size (adjusted Cohen’s *d* =1.76 vs. 1.28-1.61 for all others; Fig. 3C). However, there was no significant difference in classification accuracy despite numerically higher AUCs for BD-p-tau217%. The AUC for BD-p-tau217% for distinguishing T+ from T-individuals was 0.90(95%CI=0.81–0.95) compared to 0.88(95%CI=0.78–94) for BD-p-tau217, 0.84(95%CI=0.72–0.92) for p-tau217, 0.84(95%CI=0.74–0.91) for p-tau217%, and 0.81(95%CI=0.70–0.89) for p-tau217/Aβ42 (Fig. 3C-D and Data S2-3).

### Relation to clinical diagnosis of AD vs. non-AD in Pitt-ADRC clinical cohort

For differentiating AD dementia from CU, MCI and other neurodegenerative diseases (i.e., non-AD dementia), plasma BD-p-tau217% was the most promising across all biomarkers, had the greatest effect sizes, and the highest AUCs (Fig. 4 and Data S2-3). For instance, the AUC of plasma BD-p-tau217% for distinguishing AD dementia from CU was 0.87(95%CI=0.84–0.89; 82% correctly classified) which significantly outperformed BD-p-tau217 (0.85[95%CI=0.83–0.88]; p=0.014), p-tau217 (0.81[95%CI=0.78– 0.84]; p<0.001), p-tau217% (0.84[95%CI=0.81–0.87]; p=0.003) and p-tau217/Aβ42 (0.79[95%CI=0.75–0.81]; p<0.001).

**Figure 4.**
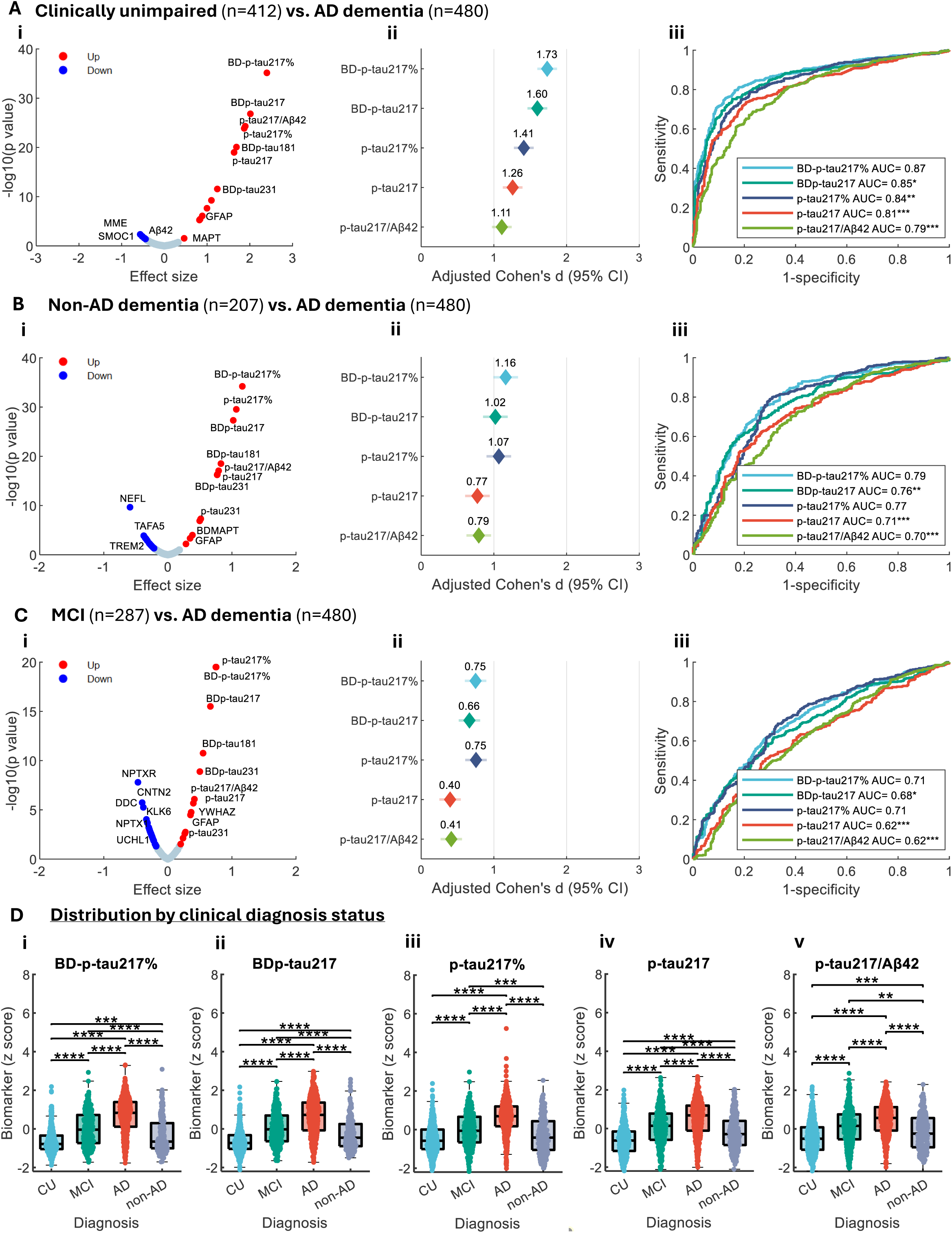
Association of plasma biomarker levels with clinical diagnosis in the Pitt-ADRC clinical cohort. Panels A-C show comparison results between control vs. AD, non-AD dementia vs. AD dementia, and MCI vs. AD, respectively. Volcano plots (Ai to Ci) display age and sex adjusted Cohen’s d effect sizes on the x axis and −log (p values) on the y axis. Red dots indicate biomarkers up regulated, and blue dots indicate biomarkers downregulated, in AD dementia. P values were FDR adjusted using the Benjamini–Hochberg procedure. Effect size forest plots (Aii to Cii) show point estimates and 95% confidence intervals of Cohen’s *d*. Biomarker levels were z normalized. AUC plots (Aiii to Ciii) illustrate logistic regression classification performance using biomarker level alone. Asterisks denote DeLong test comparisons using BD-p tau217% as the reference biomarker, with *, **, and *** indicating p < 0.05, 0.01, and 0.001, respectively. Panels Di-v display box plot distributions of biomarker levels by clinical diagnosis status. The center line indicates the median, box edges correspond to the 25th and 75th percentiles, and whiskers extend to the most extreme non outlier values. All pairwise rank sum tests yielded Benjamini– Hochberg–corrected p < 0.0001, denoted by ****.

In discriminating AD dementia vs. non-AD dementias, plasma BD-p-tau217% had an AUC of 0.79(95%CI=0.75–0.83; 74% correctly classified) which was significantly superior to those of BD-p-tau217, p-tau217/Aβ42, and p-tau217 with classification accuracies of 70%, 71%, and 66% respectively, and numerically higher than that of p-tau217% (0.77[95%CI=0.73-0.80]; Fig. 4B and Data S2-3). In separating MCI and AD dementia, BD p tau217% and p-tau217% achieved a similar effect size of 0.75 and AUC of 0.71, also significantly higher than those of p tau217, p-tau217/Aβ42, and BD-p-tau217 (Fig. 4C and Data S2-3). Figure 4D shows boxplots of plasma p-tau levels according to clinical diagnosis.

### Improved discriminative accuracies in individuals with comorbidities in Pitt-ADC clinical cohort

In the clinical cohort, we evaluated diagnostic accuracies of the plasma biomarkers in discriminating clinically classified AD dementia from CUs with six common comorbidities, including diabetes, obesity (body mass index >=30), any cardiovascular disease (CVD), hypercholesterolemia and active depression, based on information collected from health information interviews (see Materials and Methods).

Among participants whose health information indicated an absence of the respective comorbidities (i.e., without comorbidity), plasma BD p tau217% consistently maintained a higher diagnostic accuracy, with significantly greater performance than p tau217 and p tau217/Aβ42 across all sub analyses, mirroring the superiority observed in the full cohort (Fig. 5). The AUCs for BD-p-tau217% ranged between 0.87 and 0.90, compared with smaller AUCs between 0.80 and 0.83 for both p-tau217 and p-tau217/Aβ42, and 0.84 to 0.88 for p-tau217%.

**Figure 5.**
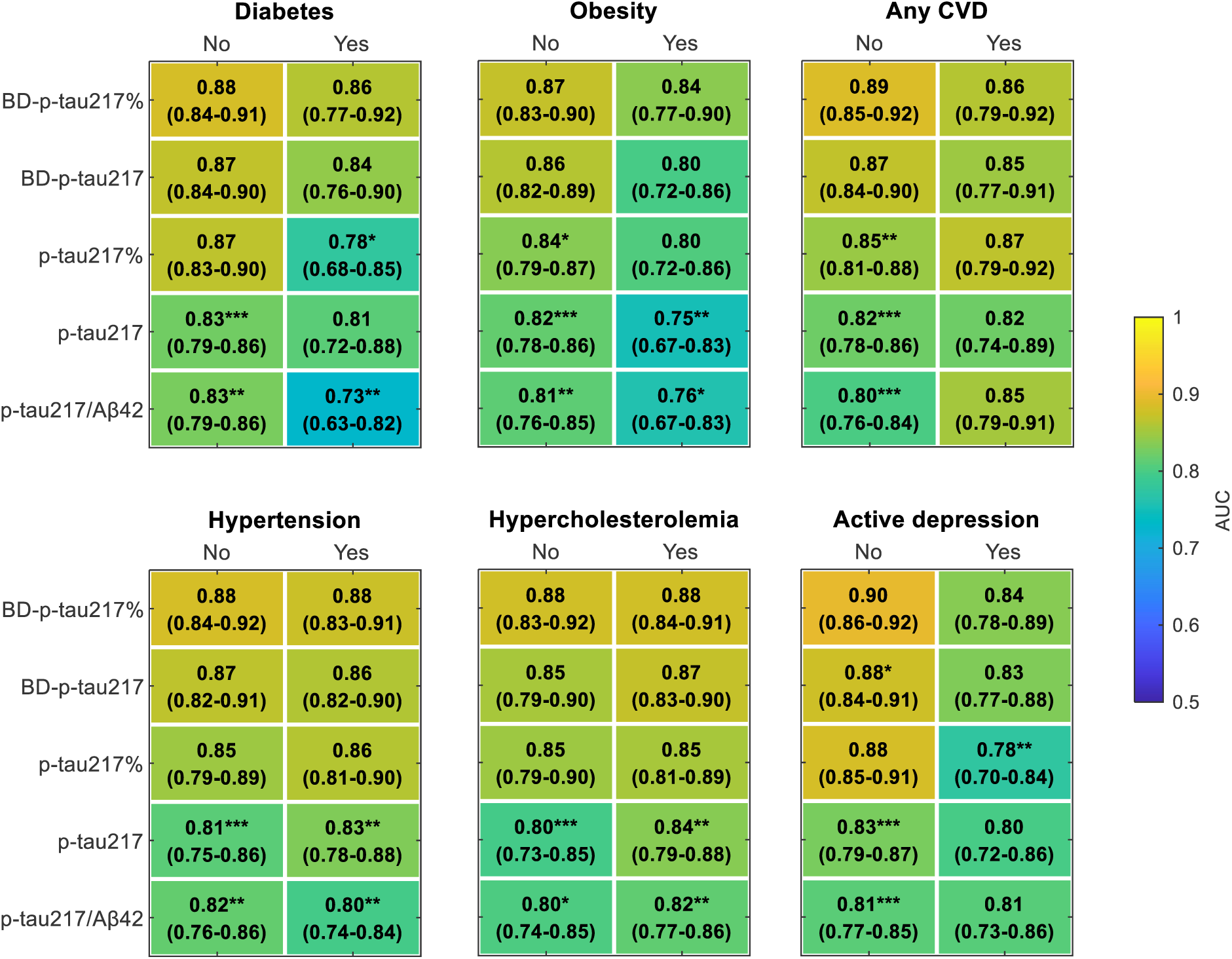
Impact of comorbidities on plasma biomarker diagnostic performance in the Pitt-ADRC clinical cohort. No and Yes indicate the absence and presence of the corresponding comorbidity, respectively. Cell annotations show AUC estimates with 95% confidence intervals for discriminating controls from individuals with AD. Asterisks denote DeLong test comparisons using BD-p-tau217% as the reference biomarker, with *, **, and *** indicating p < 0.05, 0.01, and 0.001, respectively.

In all sub-analysis focusing on participants with the indicated comorbidities, BD-p-tau217% maintained higher AUCs in all conditions of interest (except for “any CVD” where all AUCs were statistically indifferent). BD-p-tau217% maintained relatively stable AUCs in the presence of comorbidities, with AUCs ranging from 0.84 to 0.88. In participants <u>with</u> comorbid obesity, hypertension, and hypercholesterolemia, BD-p-tau217% and BD-p-tau217 remained significantly more accurate than p-tau217 and p-tau217/Aβ42 in identifying AD dementia vs. CUs (Fig. 5). The AUCs for BD p tau217% were 0.84(95%CI=0.77–0.90) for obesity, 0.88(95%CI=0.83–0.91) for hypertension, and 0.88(95%CI=0.84–0.91) for hypercholesterolemia. In contrast, the corresponding AUCs for p tau217 were 0.75(95%CI=0.67–0.83), 0.83(95%CI=0.78–0.88) and 0.84(95%CI=0.79–0.88). For p-tau217/Aβ42, they were 0.76(95%CI=0.67–0.83), 0.80(95%CI=0.74–0.84) and 0.82(95%CI=0.77–0.86).

Focusing on participants with comorbid diabetes and active depression, BD-p-tau217% showed significantly better performance than p-tau217%. Notably, p-tau217% showed relatively large (∼10%) reductions in AUC values among participants with vs. without diabetes and active depression, resulting in significantly inferior accuracies compared with BD p tau217%. For p-tau217%, the AUC values decreased sharply from 0.86(95%CI=0.77–0.92) to 0.78(95%CI=0.68–0.85) for those without vs. with diabetes, respectively, and from 0.88(95%CI=0.85-0.91) to 0.78(95%CI=0.70-0.84) when considering participants without vs. with active depression (Fig. 5).

### Prediction of future clinical progression

We next evaluated performance in predicting future diagnostic progression among individuals who were CU or had MCI at baseline (participant characteristics in Table S3). We defined optimal thresholds for each biomarker by contrasting A+ and A-participants in the neuroimaging cohort (Data S2) and compared clinical progression trajectories in individuals with high vs. low levels (Fig. 6). Dichotomization by plasma BD-p-tau217% resulted in the clearest between-group separation. In CU, the median survival times (MSTs) were 8.0 vs. 19.0 years (ΔMST=11), and in those with MCI, 3.1 vs. 12.0 years (ΔMST=8.9) for individuals with elevated vs. normal BD-p-tau217% values, respectively. BD p tau217 showed similarly robust discrimination in MCI but less so in CU (corresponding ΔMSTs of 8.7 in CU and 8.9 in MCI). In contrast, non-CNS-selective p tau217 based biomarkers exhibited weaker separation. The ΔMSTs for p tau217 were 6.7 in CU and 7.9 in MCI. For p tau217%, the ΔMSTs were 5.9 and 4.9, and for p tau217/Aβ42, 5.1 and 7.3 (Fig. 6A-B).

**Figure 6.**
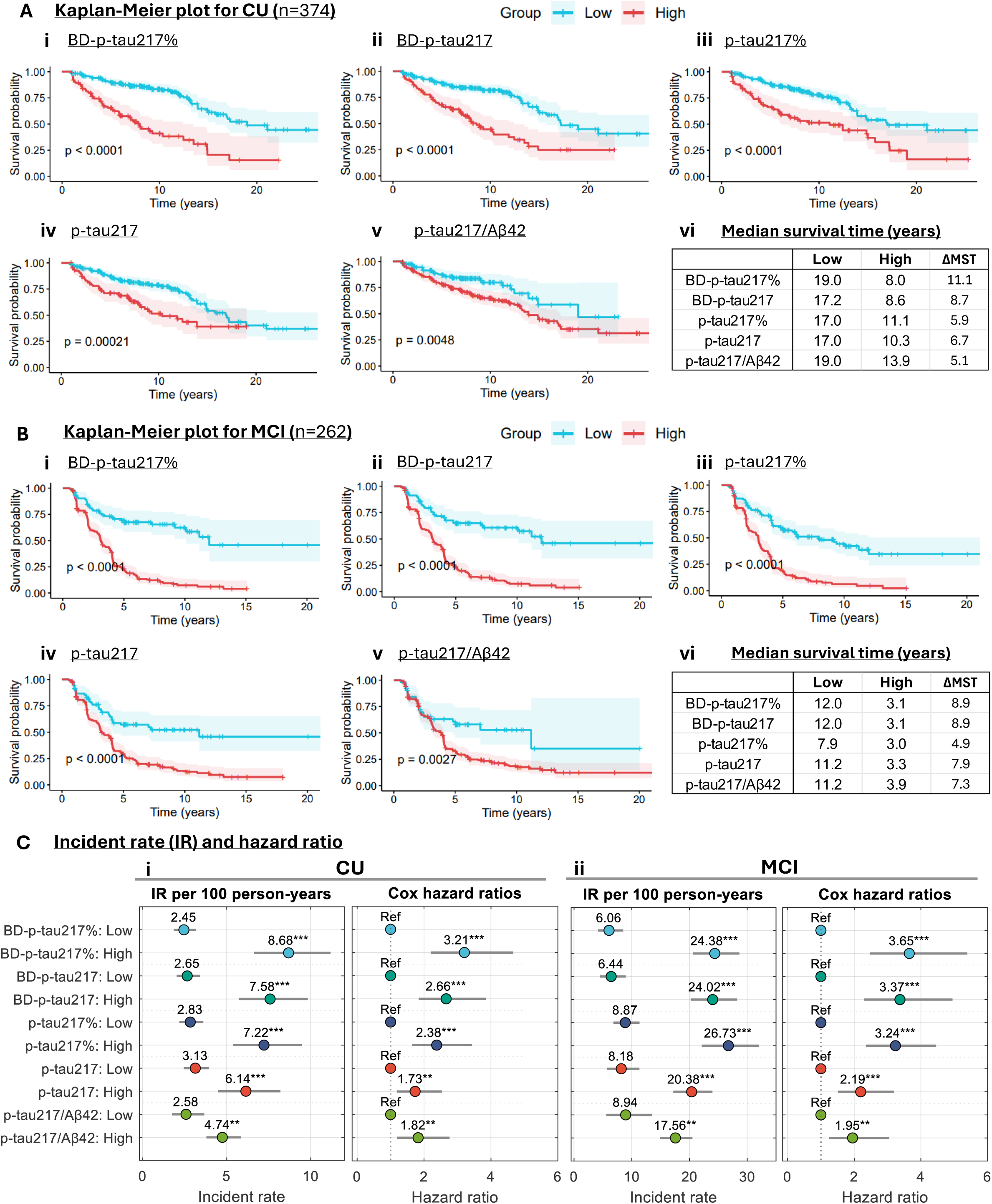
Plasma biomarker performance in predicting future cognitive decline in the Pitt-ADRC clinical cohort. Panels A and B show Kaplan-Meier survival curves illustrating time to incident clinical progression for individuals who were clinically unimpaired (A) or had MCI (B) at baseline, stratified by optimal threshold derived from differentiating Aβ-PET positivity in the neuroimaging cohort. Shaded area indicated 95% confident interval. Panel C presents forest plots of incidence rates (IRs) per 100 person years and hazard ratios (HRs) for baseline controls (Ci) and for individuals with MCI (Cii). HRs were estimated using Cox proportional hazards models adjusted for age and sex, with the low biomarker group as the reference. Asterisks in panel C indicate significance, with *, **, and *** corresponding to p < 0.05, 0.01, and 0.001, respectively. MST denotes median survival (stable) time.

Consistently, dichotomizing participants into high versus low plasma BD p tau217% most effectively separated individuals by their incidence rate (IR) and Cox hazard ratio (HR) for clinical progression (Fig. 6C). High BD p tau217% was associated with the greatest risk elevation in both CU and MCI groups, with HRs of 3.21 (95%CI=2.20–4.66) and 3.65 (95%CI=2.47–5.39), respectively, compared with the corresponding low BD p tau217% groups. Dichotomization using BD p tau217% also produced the largest separation in IRs, with 8.68 versus 2.45 per 100 person years (ΔIR=6.23) in CU, and 24.38 versus 6.06 per 100 person years (ΔIR=18.32) in MCI.

When categorized according to plasma biomarker quartiles, the MST for plasma BD-p-tau217% showed a more consistent stepwise reduction across quartiles than the other biomarkers in CU, with much clearer doubling patterns when comparing quartile-1 with quartile-2 and so on (Fig. S4A). Stratification by BD p tau217% quartiles also produced the largest overall separation in IR and HR values in CU individuals (Fig. S5A). For example, the HR for BD p tau217% quartile 4 relative to quartile 1 was 8.17 (95%CI=4.16–16.06), higher than the corresponding HRs of 5.17 (95%CI=2.69–9.93) for BD-p-tau217, 3.87 (95%CI=2.02–7.40) for p-tau217%, 2.99 (95%CI=1.54–5.82) for p-tau217, and 2.24 (95%CI=1.21–4.14) for p tau217/Aβ42. Among individuals with MCI, BD p tau217% also provided clearer risk stratification across quartiles than BD p tau217, p tau217, and p tau217/Aβ42. P tau217% showed slightly greater quartile based separation in IR and HR than BD p tau217% (Fig. S4B and S5B).

When analyzing clinical progression by progression status (progressor vs. stable; participant characteristics in Table S8), plasma BD-p-tau217% emerged as the strongest predictor of 5-year progression in both CU (progressing to MCI or AD dementia) and individuals with MCI (progressing to AD dementia; Fig. S6). BD-p-tau217% had adjusted Cohen’s *d* effect sizes of 1.39 and 0.92 for MCI and CU respectively. In contrast, p-tau217 managed only 0.69 and 0.59. In agreement, BD-p-tau217% recorded the highest AUC of 0.83 (95%CI=0.80–0.85) to distinguish progressor MCI from stable MCI, which was significantly superior to 0.78 (95%CI=0.75–0.80; p=0.013) for BD-p-tau217, 0.69 (95%CI=0.66–0.72; p<0.001) for p-tau217 and 0.65 (95%CI=0.62–0.68; p<0.001) for p-tau217/Aβ42 (Fig. S6A and Data S2-3). In terms of separating progressor controls from stable controls, we found no statistical difference in the AUC values despite that of BD-p-tau217% being numerically higher than the rest (Fig. S6B). Yet, progressor controls had significantly higher BD-p-tau217% and other p-tau217 biomarker levels than stable controls, just as progressor MCIs had higher levels than stable MCIs (Fig. S6C).

### Sensitivity analyses

Statistical details of all biomarker performance comparisons beyond AUCs, including accuracy, sensitivity, specificity, positive (PPV) and negative predictive values (NPV) are shown in Data S2. DeLong tests for the significance of AUC comparisons are provided in Data S3. Risk tables of all survival analysis were available in Data S4. In addition to the AUCs and effect size findings described above, plasma BD-p-tau217% maintained superiority over p-tau217 and p-tau217/Aβ42 (and in several key comparisons p-tau217%) in nearly all primary and secondary analyses when comparing other important metrics such as sensitivity, specificity, accuracy, NPV and PPV, further reinforcing the strong diagnostic value of BD-p-tau217% (Data S2).

All AUC values described above were derived from biomarker only models. AUCs from models additionally adjusted for age and sex are provided in Data S5 and show trends consistent with those observed in the biomarker only analyses. All effect sizes reported above were adjusted for age and sex, demographic factors known to influence AD blood biomarker levels. As a sensitivity analysis, we also evaluated effect sizes for the same comparisons using biomarker only models, as well as models adjusted for a broader set of covariates (age, sex, race, education, and *APOE4* carrier status; Data S6). Under both modeling strategies, BD p tau217% consistently demonstrated the largest effect sizes across nearly all comparisons, ranging from 0.75 to 2.55 in biomarker only models and from 0.66 to 2.72 in the fully adjusted models. In contrast, the corresponding effect size ranges were 0.40–2.04 and 0.39–2.10 for p tau217, 0.40–2.07 and 0.35–2.10 for p tau217/Aβ42, 0.66–2.23 and 0.54–2.38 for BD p tau217, and 0.57–1.89 and 0.48–2.01 for p-tau217%.

### External validation in the ADNI cohort

Results in the ADNI cohort mirrored those in the Pitt-ADRC cohorts. BD-p-tau217% outperformed all other NULISA biomarkers in detecting abnormal Aβ-PET and tau-PET and in discriminating CU from AD (Fig. 7). For discriminating A+ from A− participants, BD-p-tau217% achieved the largest effect size of 2.20, followed by 1.94 for BD-p-tau217, 1.87 for p-tau217/Aβ42, 1.77 for p-tau217%, and 1.76 for p-tau217 (Fig. 7A). BD-p-tau217% also achieved the highest AUC (0.93; 95% CI =0.91-0.95), significantly exceeding those of BD-p-tau217 (0.92; 95% CI=0.90-0.94; p = 0.013), p-tau217/Aβ42 (0.91; 95% CI=0.89-0.93; p = 0.007), p-tau217 (0.90; 95% CI=0.88-0.92; p < 0.001), and p-tau217% (0.89; 95% CI=0.87-0.91; p < 0.001). For detecting tau-PET positivity, BD-p-tau217% achieved an effect size of 2.27, closely followed by 2.22 for BD-p-tau217 (Fig. 7B). Both biomarkers achieved similar AUCs of 0.92, significantly higher than that of p-tau217 (0.89 [95%CI=0.84-0.92]; p = 0.029 and p = 0.012 for BD-p-tau217% and BD-p-tau217, respectively). In addition, the AUC of BD-p-tau217% was significantly higher than that of p-tau217/Aβ42 (0.88 [95%CI=0.81-0.92]; p = 0.046).

**Figure 7.**
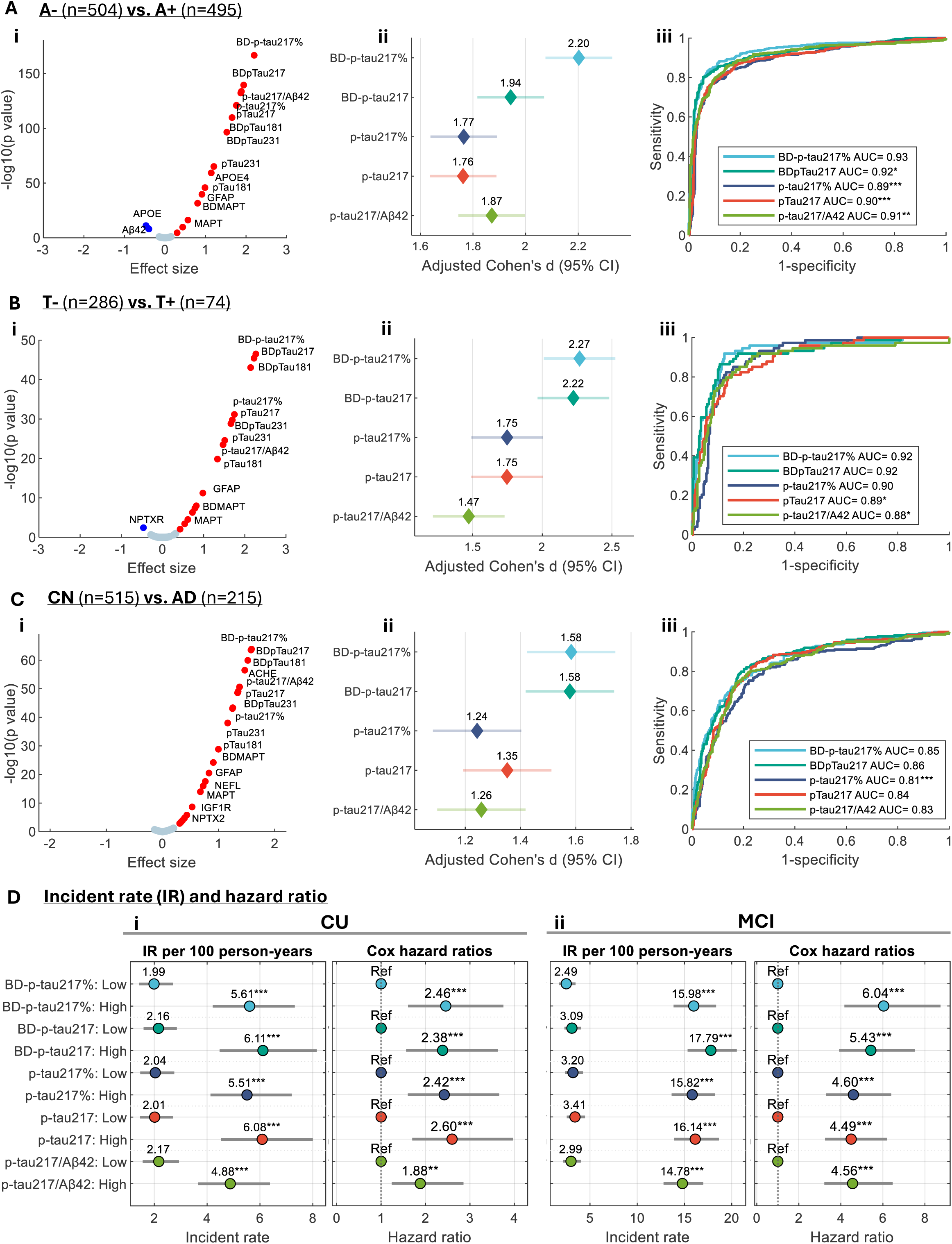
External validation in the ADNI cohort. (A-C) Biomarker performance for discriminating A+ versus A−, T+ versus T−, and cognitively unimpaired (CU) versus AD participants, respectively. Volcano plots (Ai-Ci) show age- and sex-adjusted Cohen’s d effect sizes versus −log (p), with p values adjusted using the Benjamini-Hochberg procedure. Effect size forest plots (Aii-Cii) display adjusted Cohen’s d estimates and 95% confidence intervals. Biomarker levels were z-normalized. AUC plots (Aiii-Ciii) show logistic regression classification performance based on biomarker levels alone. Asterisks denote DeLong test comparisons against BD-p-tau217%. (D) Incidence rates (IRs) per 100 person-years and hazard ratios (HRs) for clinical progression among baseline CU (Di) and MCI (Dii) participants. HRs were estimated using age- and sex-adjusted Cox proportional hazards models, with the low biomarker group as the reference. *, **, and *** indicate p < 0.05, 0.01, and 0.001, respectively.

In discriminating CU from AD, BD-p-tau217% and BD-p-tau217 achieved the largest effect sizes (both 1.58) and highest AUCs (both 0.92), compared with effect sizes of 1.24-1.35 and AUCs of 0.88-0.90 for p-tau217-based measures (Fig. 7C). The AUCs of both BD-p-tau217% and BD-p-tau217 were significantly higher than that of p-tau217% (p<0.001). Similar superiority of BD-p-tau217-based measures over p-tau217-based analytes were observed for discriminating MCI from CU and AD from MCI (Fig. S7).

When dichotomized into high- and low-biomarker groups, all five biomarkers showed similar performances for predicting clinical progression among baseline CU participants, with HRs ranging from 2.38 to 2.60 (Fig. 7Di). In contrast, among baseline MCI participants, BD-p-tau217% and BD-p-tau217 provided greater separation of progression risk, with ΔIRs of 13.49 and 14.70 per 100 person-years and HRs of 6.04 and 5.43, respectively, compared with ΔIRs <12 and HRs of 4.49-4.60 for the other biomarkers (Fig. 7Dii).

### Evaluation of alternative ratio-based biomarkers

We next evaluated whether alternative ratio-based biomarkers provided improvements similar to those observed for BD-p-tau217% relative to BD-p-tau217. Figure S8 summarizes the effect sizes across neuropathological, PET, and clinical outcomes for combinations of BD-p-tau217 or p-tau217 normalized to BD-tau, total tau, Aβ42, and Aβ40. Among all ratios examined, only BD-p-tau217/BD-tau (BD-p-tau217%) consistently improved performance over the corresponding single biomarker across both the Pitt-ADRC and ADNI cohorts. In contrast, normalization of BD-p-tau217 to total tau, Aβ42, or Aβ40 generally reduced performance, with the largest decline observed for Aβ40, followed by total tau and Aβ42. Normalization of p-tau217 to Aβ42 modestly improved the discrimination of low versus severe ADNC but did not consistently enhance the detection of Aβ-PET or tau-PET positivity or the discrimination of clinical diagnostic groups across cohorts (Figure S8).

## DISCUSSION

We have shown that selective quantification of brain-derived p-tau217 substantially improves the diagnostic and prognostic performance of plasma p-tau217 using data from independent cohorts. Plasma BD-p-tau217% and BD-p-tau217 were significantly more accurate than standard p-tau217-based biomarkers, including p-tau217 alone, p-tau217% and the p-tau217/Aβ42 ratio as biomarkers for AD. These superior accuracies were observed across neuropathologically-verified ADNC, amyloid-PET, tau-PET, clinical diagnostic, and longitudinal progression outcomes. Furthermore, the plasma BD-p-tau217 versions remained the best-performing biomarker with the largest accuracies to detect AD in older adults with common comorbidities. These findings demonstrate that BD-p-tau217% and BD-p-tau217 capture AD pathological signatures with greater specificity and sensitivity and are much less prone to unwanted impacts from comorbid conditions.

This work introduces a conceptual advance: molecular source specificity as a principle for blood-based biomarker development. Instead of measuring total circulating analyte concentration alone (as is being done for current p-tau217-based methods), enriching for predominantly CNS-derived pools can substantially enhance disease relevance and robustness. This is the most comprehensive study yet on BD-p-tau, across multiple prognostic, diagnosis and longitudinal angles, extending recent studies limited to Aβ-PET-characterized cohorts (*47, 48*). Moreover, it is the first to introduce the BD-p-tau217% concept, building on its early adoption for standard p-tau217 (*46*).

While both BD-p-tau217% and BD-p-tau217 outperformed conventional p-tau217 and p-tau217-based biomarkers in most analyses, the former had a slight advantage for the following reasons: (1) the ratio appeared less prone to comorbidity effects; (2) BD-p-tau217% had numerically higher AUCs, accuracies, sensitivity, specificity, NPV and PPV despite these not always being significant; (3) BD-p-tau217% had larger effect sizes, hazard ratios and incident rates; and (4) BD-tau – and for that matter – BD-p-tau217% is less prone to effects of variables such as age and sex(*45*). Interestingly, among all ratio-based biomarkers evaluated (BD-p-tau217 and p-tau217 normalized to BD-tau, total tau, Aβ40, or Aβ42), only BD-p-tau217% consistently improved performance across both the Pitt-ADRC and ADNI cohorts. Biologically, BD-p-tau217 reflects the abundance of CNS-enriched tau phosphorylated at threonine 217, whereas BD-p-tau217% may more closely approximate phosphorylation occupancy by accounting for its corresponding tau substrate. The improved performance of BD-p-tau217% suggests that phosphorylation occupancy may provide additional pathological information beyond absolute p-tau217 levels, particularly during the early stages of AD.

It is worth noting that while p-tau217% showed modest improvements over p-tau217 in the Pitt-ADRC cohorts for distinguishing low or intermediate from severe ADNC and for predicting clinical diagnosis and progression, these improvements were not replicated in the ADNI cohort. P-tau217% was originally developed in the PrecivityAD2 mass spectrometry assay, where p-tau217 is normalized to its non-phosphorylated counterpart (np-tau217) and this normalization was shown to improve performance relative to p-tau217 alone (*46, 49, 50*). Unlike p-tau217% in the PrecivityAD2 assay, which normalizes p-tau217 to non-phosphorylated (np)-tau217, our implementation used total tau as the denominator since np-tau217 is not measured in the NULISA panel. Whether differences in normalization strategy contributed to the discrepant findings remains unclear and warrants further investigation. It will also be of interest to determine whether normalizing mass spectrometry–based p-tau217 to BD-tau, quantified using peptides spanning the exon 4/5 junction, provides greater diagnostic utility than normalization to np-tau217.

The advantages of BD-p-tau217% and BD-p-tau217 over traditional p-tau217 based measures were mainly observed in: (1) detecting Aβ-PET positivity but not tau PET abnormalities; (2) distinguishing low from intermediate as well as intermediate vs. high ADNC stages, but not low and high ADNC groups; (3) tracking longitudinal cognitive decline; and (4) in individuals with comorbidities. This indicates that BD-p-tau-based biomarkers are superior for detecting early, emerging AD with incipient Aβ pathology when tau NFTs are not yet highly accumulated. What is the clinical relevance of these findings? The closer a blood biomarker mirrors the intended features of a standard of truth measure, the better its performance in linking clinical symptoms to the underlying disease biology(*51*). Plasma p-tau217 holds a crucial role in accessible AD diagnostics and care(*17, 52, 53*). Our results show BD-p-tau217% and BD-p-tau217 as substantive next generation biomarkers with enhanced performance to transform the detection, treatment and monitoring of AD.

For more direct head-to-head comparisons, we focused on biomarkers measured using the same NULISA proteomic panel, sharing identical p-tau217 antibodies, buffer systems, and platform sensitivity, therefore minimizing variability related to sample processing, batch effects, and cross-platform analytical differences. Previous head-to-head comparison studies have reported that NULISA performs comparably to other leading immunoassay-based platforms (*54–56*). Although it was not the top-performing platform in a large round-robin study (*56*), it offers high analytical sensitivity and the ability to simultaneously quantify many low-abundance proteins from a small plasma volume (*43*). Importantly, BD-p-tau217% achieved AUCs for detecting Aβ positivity comparable to those reported for mass spectrometry-based plasma p-tau217/np-tau217 assays. This level of performance is notable given that mass spectrometry is generally considered the benchmark platform yet requires substantially greater analytical complexity and sample volume.

The enhanced performance of BD-p-tau217% and BD-p-tau217 has several potential clinical implications. Current AD diagnostic guidelines recommend plasma p-tau217 testing as part of clinical decision making in specialist memory clinics (*8, 18*). The superior diagnostic accuracy of BD-p-tau217% and BD-p-tau217, particularly in detecting Aβ pathology, suggest that these biomarkers may be better suited for triaging and confirmatory diagnosis. This may be especially relevant in the context of anti-Aβ therapeutics, which requires demonstration of Aβ pathology among cognitively impaired individuals (*57, 58*). Other relevant situations include enrolment and monitoring in therapeutic trials as well as in primary care- and population-based screening programs.

The improved prognostic performance further enhances the clinical value of these BD-p-tau biomarkers. BD-p-tau217% was the most accurate for identifying MCI individuals who will progress to AD dementia within 5 years in the Pitt-ADRC cohort and provided the greatest separation of incident AD risk in both the Pitt-ADRC and ADNI cohorts. Among MCI individuals eligible for anti-Aβ therapy, this increased predictive capacity may be crucial in classifying who should be prioritized for treatment based on their projected disease course. In addition, the stronger accuracies of BD-p-tau217 forms in individuals with comorbidities is an attractive attribute that addresses a well-known challenge for p-tau217(*26*), with similar performances in people without those comorbidities providing proof-of-concept for the versatility of BD-p-tau217%. By removing the negative impacts of comorbidities on p-tau217, plasma BD-p-tau217% may enable more widespread applications of blood-based molecular testing irrespective of comorbidity burden.

Despite the strong overall performance of all biomarkers evaluated, particularly for detecting Aβ pathology, we observed a subset of individuals with low biomarker levels despite high pathology. The causes of this apparent discordance remain unclear, as we found no consistent associations with the interval between blood collection and pathological assessment, clinical progression, or demographic characteristics. Notably, the discordance appeared more pronounced for ratio-based biomarkers, likely reflecting the additional variability introduced by both the numerator and denominator measurements. The complex biological origins of tau may also contribute to the observed discordance. Although the LMW tau species measured by the BD-tau and BD-p-tau assays are enriched in the CNS, their expression has also been reported in the PNS, where it may account for up to 50% of total tau (*38*). Likewise big tau is primarily expressed in PNS, but its expression has also been found in selected brain regions such as the cerebellum and cranial nerve motor nuclei, as well as non-neuronal cells (*42, 59*). These findings highlight the complexity of tau biology and warrant the need for additional studies to better understand the sources of biomarker-pathology discordance. This study has several strengths, including inclusion of cohorts spanning neuropathological diagnosis, molecular imaging of Aβ and tau, clinical diagnosis, and real-world electronic health record-based comorbidity assessments – enabling in-depth clinical-biological evaluation. We further validated our findings from a large academic memory clinic cohort enriched for individuals with cognitive concerns in the independent ADNI cohort, a highly characterized research cohort, demonstrating the reproducibility and generalizability of the observed associations.

A few limitations should be noted. First, although we included individuals from independent studies, they mostly included non-Hispanic White participants, and it remains unclear whether these findings generalize to other racial and ethnic populations. Second, while the NULISA proteomic panel enables simultaneous measurement of a large number of low-abundance proteins from a small plasma volume, the current assay provides relative rather than absolute quantification, which may limit direct clinical implementation. However, single-analyte biomarker test kits for BD-p-tau217 and BD-tau potentially suitable for *in vitro* diagnostic tests or laboratory developed tests have been developed(*47, 60, 61*). Future work will aim toward clinical translation of these tests to support clinical decision making.

Together, plasma BD-p-tau217% and BD-p-tau217 capture AD pathology and clinical progression prediction with significantly superior performances – including AUCs, accuracies, effect sizes, sensitivity and specificity –over conventional, non-CNS-selective p-tau217 biomarkers. These findings were verified in cohorts with paired antemortem blood and postmortem neuropathology assessments, molecular imaging of brain Aβ and tau, and in memory clinic participants with up to two decades of clinical monitoring. These results show that BD-p-tau217% has the potential to address key limitations of existing p-tau217 assays while further transforming their clinical performances. Future studies should evaluate longitudinal biomarker dynamics, and implementation in population studies and in primary care. Yet, the breadth and consistency of evidence presented here support BD-p-tau217% as an important biomarker for the next generation of precision AD diagnosis and prognosis that will minimize false positives and maximize diagnostic and prognostic utility.

## MATERIALS AND METHODS

### Study design & population

The Pitt□ADRC cohort comprises adults self-or provider-referred for cognitive evaluation, volunteering for research, and/or recruited via educational outreach events largely from the greater southwestern Pennsylvania area, enrolled in a registry to study Alzheimer disease, aging and related disorders. Inclusion criteria consist of individuals aged 60 years or older, as well as younger adults who self□report symptoms suggestive of Alzheimer disease and related dementias, and availability of reliable study partner/informant. Exclusion criteria include significant psychiatric conditions or other medical disorders that could interfere with the accurate assessment of cognitive impairment, and history of cancer (other than skin and in situ prostate) within the past 5 years. Detailed sociodemographic data, such as age, sex, self□reported race, and educational background, were obtained at each participant’s baseline visit. Participants also completed an extensive set of baseline evaluations, including questionnaires assessing overall cognitive, behavioral, emotional, and medical status, a comprehensive battery of clinical assessments, and provided blood for biomarker measurement and genotyping. Following baseline, individuals returned annually for clinical follow□up evaluations. The Pitt□ADRC study was approved by the University of Pittsburgh Institutional Review Board (STUDY19110245), and all participants provided written informed consent.

This study included a subset of participants who were enrolled in the Pitt□ADRC between 1993 and 2023. Participants were selected based on the availability of plasma samples and were prioritized if they had available AD pathology from post□mortem neuropathology (death within 10 years from blood draw), antemortem neuroimaging (within two years from blood draw), or longitudinal clinical follow□up. Because all participants were drawn from the Pitt□ADRC, some individuals appear in more than one of the analytic cohorts (Fig. S2).

The ADNI study was launched in 2004 as a public-private partnership led by Principal Investigator Michael W. Weiner, MD, with the goal of characterizing longitudinal changes in cognition, function, neuroimaging measures, and fluid biomarkers in cognitively unimpaired individuals, participants with MCI, and those with AD. ADNI data used in the preparation of this article were obtained from the ADNI website (https://adni.loni.usc.edu), downloaded in May 2026. Detailed study design has been previously described (*62, 63*).

### Clinical assessments for the Pitt-ADRC

A detailed description of the clinical assessments, including neurologic examination, neuropsychological assessment, psychiatric evaluation and diagnostic tests such as brain magnetic resonance imaging (MRI), has been published previously(*64*). Clinical diagnoses were assigned at baseline and at each annual follow□up during a clinical consensus conference, in which the study team (comprising neurologists, neuropsychologists, psychiatrists, and related dementia care specialists) reviewed all available clinical data, laboratory results, and MRI findings based on published criteria. In this study, participants were classified as clinically unimpaired (CU; include both cognitively normal and subjective complaint with normal test score), MCI, AD dementia (both probable and possible AD), or non□AD dementia. For MCI, we excluded individuals with non□amnestic presentations.

We classified participants who were CU or had MCI at baseline as either stable or progressors based on their clinical trajectory over a 5□year period. Baseline CU who progressed to MCI or AD dementia within five years were classified as progressor CU, whereas those who remained CU at their first visit after the 5□year interval were classified as stable CU. Similarly, baseline MCI participants who progressed to AD dementia were classified as progressor MCI, while those who remained MCI or reverted to CU at the first post□5□year visit were classified as stable MCI.

Medical history was extracted from participants’ health history questionnaires. For this study, we focused on six common comorbidities: diabetes, obesity, any cardiovascular disease (defined as having at least one of the following conditions or histories—angina, myocardial infarction, atrial fibrillation, congestive heart failure, prior heart attack, angioplasty, cardiac bypass, pacemaker implantation, heart valve replacement, or other cardiovascular diseases), hypertension, hypercholesterolemia, and active depression. For all comorbidities, we categorized individuals as either having no history of the condition or having a recent/active condition, defined as occurring within the past year (except for depression which was classified based on occurrences within the past two years) or still requiring ongoing clinical management. Remote or resolved conditions, those that occurred in the past and no longer required treatment, were considered missing and excluded from statistical analyses.

### Pitt-ADRC neuropathological assessment

The neuropathological data was collected following the National Alzheimer’s Coordinating Center (NACC) data collection procedures (versions 1, 7, 9, 10, and 11)(*65*). Due to missing Thal phase for a subset of cases, we implemented a modified NIA-Reagan approach for classifying ADNC(*66*). Neuritic plaque burden was assessed using the CERAD score for neocortical neuritic plaque density(*67*), where scores of 0–1 indicate low, 2 indicates moderate, and 3 indicates frequent plaque density. NFT pathology was classified into four groups using the Braak staging system(*68*): Braak stages 0–II were considered low□Braak; stages III–IV with no or sparse neuritic plaques were classified as primary age□related tauopathy (PART); stages III–IV with moderate or greater neuritic plaque density were considered mid□Braak; and stages V–VI were categorized as high□Braak. ADNC was defined as followed: intermediate ADNC was Braak III-IV with moderate/frequent CERAD neuritic plaque score, and high ADNC was Braak V-VI with moderate/frequent CERAD neuritic plaque score. Everyone else was considered Low/Not ADNC. In the neuropathology cohort, all individuals with intermediate ADNC belonged to the mid□Braak group and vice versa, and all individuals with severe ADNC had high□Braak and vice versa. Therefore, comparisons between mid Braak and high Braak groups were not included in the analysis.

### Amyloid and tau PET imaging in Pitt-ADRC

All PET imaging scans for Pitt-ADRC were acquired similar as described (*69–71*). [¹¹C]Pittsburgh Compound□B ([¹¹C]PiB) was used as the tracer for Aβ□PET, and [¹□F]AV□1451 was used for tau□PET. Participants received each tracer via slow bolus injection through the antecubital vein, followed by a brief resting period before being positioned in the scanner for image acquisition. [¹¹C]PiB scans were obtained 50– 70 minutes post injection, and [¹□F]AV□1451 PET scans were obtained 80–100 minutes post□injection, each acquired as a series of 5 minute frames. For Aβ□PET, scans were processed using CapAIBL, a tool developed by the Australian eHealth Research Centre to simplify and standardize the quantification of amyloid burden in PET imaging(*72*). Participants were classified as A+ or A− using the CapAIBL Centiloid cutoff of 20, with values ≥20 indicating A+ status. For tau□PET, composite standardized uptake ratios (SUVRs) were calculated by normalizing the composite Braak regional values (Braak I–VI) to cerebellar gray matter activity as defined by FreeSurfer. Participants with SUVR > 1.18 were classified as T+, and those with SUVR ≤ 1.18 were classified as T−.

PET imaging for the ADNI cohort was performed as previously described (*73, 74*). Aβ-PET positivity was defined using UC Berkeley–harmonized Centiloid values, with Centiloid >20 indicating A+. Only [¹□F]AV-1451 tau-PET scans were included, with positivity defined using a previously established meta-temporal SUVR cutoff of 1.37 (*74*).

### Pitt-ADRC biospecimen collection and biomarker measurement

Blood was collected into standard ethylenediaminetetraacetic acid (EDTA) tubes at baseline visits and processed into plasma and buffy coat by centrifugation at 2,000 g for 10 minutes. *APOE* genotyping was performed as previously described(*75*). For the NULISAseq CNS panel proteomic assay, plasma samples were further centrifuged at 4,000 g for 10 minutes to remove particulates, after which 30 μL of cleared plasma was transferred to the sample plate. Assays were conducted on the Argo HT following the manufacturer’s protocol. Immunoreaction involved incubating plasma samples with a cocktail containing 132 pairs of oligo□DNA–conjugated capture and detection antibodies targeting 131 biomarkers (Data S1), along with an mCherry internal standard. Reporter DNA corresponding to each biomarker/mCherry complex was quantified using next□generation sequencing (NGS) on a NextSeq 2000 system. Reporter counts were converted to NULISA Protein Quantification (NPQ) values using Alamar Analysis software by normalizing against mCherry and inter-plate controls, followed by log2 transformation. To monitor assay performance, we included three pooled in□house QC plasma samples, run in singleton on each plate, along with vendor built-in sample control (SC), which were run in triplicate on each plate.

### Statistical analysis

Data analysis was performed using MATLAB (version R2021b). For demographic variables, the Wilcoxon rank sum test was used for two□group comparisons of continuous variables, and the Kruskal–Wallis test was used for comparisons involving more than two groups. Fisher’s exact test was applied to categorical variables with two levels, and the chi□square test was used for categorical variables with more than two levels.

Biomarker ratios were calculated using the NPQ difference. For the p□tau217/Aβ42 ratio, the value was computed as NPQ(p□tau217) − NPQ(Aβ42). For BD□p□tau217%, it was NPQ(BD□p□tau217) − NPQ(BD MAPT). For p□tau217%, the ratio was calculated as NPQ(p□tau217) − NPQ(MAPT). To enable direct comparison of effect sizes, all biomarker levels were converted to z□scores to standardize their scales.

Linear regression models were used to evaluate the association between biomarker levels and sample groupings reflecting different AD pathology or clinical diagnostic statuses. Z□score–normalized biomarker levels were used as the outcome, with sample group as the predictor, adjusting for age and sex. The adjusted Cohen’s *d* effect sizes were calculated by taking the regression coefficient for the two□category group variable (which corresponds to the adjusted group difference) and normalizing it by the residual standard deviation. For sensitivity analyses, effect sizes were also estimated using linear regression models that included either no covariates or an expanded covariate set additionally comprising race, education, and *APOE4* carriership. Multiple comparisons were controlled using the Benjamini–Hochberg procedure (1995) (*76*), and an FDR adjusted p value < 0.05 was considered statistically significant.

Receiver operating characteristic (ROC) curves and area under the curve (AUC) values were estimated using generalized linear regression models, with sample grouping as the outcome and biomarker levels as the predictor for the primary analysis. For sensitivity analyses, age and sex were added as covariates. Confidence intervals were obtained using bootstrap resampling (1,000 replicates). Classification performance was evaluated using the Youden index, which identifies the cutoff that maximizes sensitivity and specificity. Pairwise comparisons of AUCs derived from the same sample set were conducted using DeLong tests (*77*).

For longitudinal analysis, an event was defined as clinical progression from CU to MCI or AD dementia, or from MCI to AD dementia. Time□to□incident progression was calculated as the interval between the baseline blood draw and the first visit at which an event occurred, with participants who did not experience an event censored at their last completed visit. Incidence rates (IRs) were computed as the number of events divided by the total person□years of follow□up, and 95% confidence intervals were estimated using the exact Poisson method. Comparisons against the reference group (quartile-1 for quartile□based analyses or the low□biomarker group using PET□derived thresholds for dichotomized analyses) were performed using Poisson generalized linear models with log□transformed follow□up time as the offset. Cox proportional hazards models were used to evaluate the association between baseline biomarker levels and time□to□incident clinical progression among participants who were cognitively normal or had MCI at baseline, adjusting for age and sex. Kaplan-Meier models were generated to visualize survival curves and estimate median time to progression.

## List of Supplementary Materials

Figs. S1 to S8

Tables S1 to S8

Data S1 to S6

## Data Availability

The data supporting this study contain sensitive human participant information and cannot be deposited in a public repository due to IRB restrictions and the terms of participant consent. De-identified data can be shared with qualified and identifiable investigators for the purpose of replicating the results and procedures in the study. Requests can be made to the corresponding author (T.K.K.) who will direct them to the respective cohort principal investigators where necessary. Requests will be reviewed by the investigators and respective institutions to ensure that data sharing requests conform to US and EU legislation on data protection, intellectual property and confidentiality obligations. Data request to the Pittsburgh ADRC can be made directly at https://www.adrc.pitt.edu/for-researchers/adrc-data-resources/.

## Acknowledgements

The authors are thankful to the Pitt-ADRC participants and study coordinators whose dedication and commitment were fundamental to the successful implementation of this study, MIK and members of the MIK Laboratory for the storage of ADRC samples over the past 30 years.

## Author Contributions

Conceptualization: XZ, OLL, TKK; Cohort recruitment and retention: OLL, CES; Sample processing and data collection: XZ, MFF, MNN; Clinical data acquisition: BES, SBB, RAS, TAP, NKN, OLL; Data analysis: XZ, DSS, DLT, RS; PET imaging: BJL, HJA, ADC, VLV, OLL; Neuropathological assessments: DSS, JKK, MDI; Genotyping data acquisition: MIK; Manuscript draft and revision: XZ, MFF, MNN, DSS, BJL, DLT, RS, SBB, RAS, CES, BES, JKK, NKN, HJA, MDI, TAP, VLV, MIK, ADC, OLL, TKK.

## References and Notes

1. Alzheimer’s Disease International. World Alzheimer Report 2025: Reimagining life with dementia – the power of rehabilitation (Alzheimer’s Disease International, London, 2025).

2. C. R. Jack, Jr., D. M. Holtzman, Biomarker Modeling of Alzheimer’s Disease. Neuron 80, 1347– 1358 (2013).

3. R. J. Bateman et al., Clinical and biomarker changes in dominantly inherited Alzheimer’s disease. N Engl J Med 367, 795–804 (2012).

4. J. Jia et al., Biomarker Changes during 20 Years Preceding Alzheimer’s Disease. N Engl J Med 390, 712–722 (2024).

5. C. R. Jack, Jr., et al., NIA-AA Research Framework: Toward a biological definition of Alzheimer’s disease. Alzheimers Dement 14, 535–562 (2018).

6. J. Cummings, Lessons Learned from Alzheimer Disease: Clinical Trials with Negative Outcomes. Clin Transl Sci 11, 147–152 (2018).

7. T. A. Pascoal et al., Insights into the use of biomarkers in clinical trials in Alzheimer’s disease. eBioMedicine 108, (2024).

8. C. R. Jack, Jr., et al., Revised criteria for diagnosis and staging of Alzheimer’s disease: Alzheimer’s Association Workgroup. Alzheimers Dement 20, 5143–5169 (2024).

9. D. O. T. Alawode et al., Transitioning from cerebrospinal fluid to blood tests to facilitate diagnosis and disease monitoring in Alzheimer’s disease. J Intern Med 290, 583–601 (2021).

10. J. Contador, A. M. Vargas-Martínez, R. Sánchez-Valle, M. Trapero-Bertran, A. Lladó, Cost-effectiveness of Alzheimer’s disease CSF biomarkers and amyloid-PET in early-onset cognitive impairment diagnosis. European Archives of Psychiatry and Clinical Neuroscience 273, 243–252 (2023).

11. W. G. Balogun, H. Zetterberg, K. Blennow, T. K. Karikari, Plasma biomarkers for neurodegenerative disorders: ready for prime time? Curr Opin Psychiatry 36, 112–118 (2023).

12. T. K. Karikari et al., Blood phospho-tau in Alzheimer disease: analysis, interpretation, and clinical utility. Nat Rev Neurol 18, 400–418 (2022).

13. V. Pacoova Dal Maschio et al., The Role of Blood-Based Biomarkers in Transforming Alzheimer’s Disease Research and Clinical Management: A Review. International Journal of Molecular Sciences 26, 8564 (2025).

14. S. Janelidze et al., Head-to-head comparison of 10 plasma phospho-tau assays in prodromal Alzheimer’s disease. Brain 146, 1592–1601 (2023).

15. A. Silva-Spínola et al., Plasma p-tau217, quantified by the fully automated LUMIPULSE G platform, outperforms p-tau181 in predicting amyloid pathology in cognitive complaints patients. Scientific Reports 16, 4133 (2026).

16. J. Therriault et al., Blood phosphorylated tau for the diagnosis of Alzheimer’s disease: a systematic review and meta-analysis. Lancet Neurol 24, 740–752 (2025).

17. G. Grande et al., Blood-based biomarkers of Alzheimer’s disease and incident dementia in the community. Nature Medicine 31, 2027–2035 (2025).

18. S. Palmqvist et al., Alzheimer’s Association Clinical Practice Guideline on the use of blood-based biomarkers in the diagnostic workup of suspected Alzheimer’s disease within specialized care settings. Alzheimers Dement 21, e70535 (2025).

19. S. E. Schindler et al., Acceptable performance of blood biomarker tests of amyloid pathology — recommendations from the Global CEO Initiative on Alzheimer’s Disease. Nature Reviews Neurology 20, 426–439 (2024).

20. U.S. Food and Drug Administration. FDA clears first blood test used in diagnosing Alzheimer’s disease (2025), (available at https://www.fda.gov/news-events/press-announcements/fda-clears-first-blood-test-used-diagnosing-alzheimers-disease).

21. Quest Diagnostics. Quest Diagnostics launches new AD Detect blood test to aid in confirming Alzheimer’s disease (2025), (available at https://newsroom.questdiagnostics.com/2025-04-09-Quest-Diagnostics-Launches-New-AD-Detect-TM-Blood-Test-to-Aid-in-Confirming-Alzheimers-Disease).

22. C2N Diagnostics. C2N Diagnostics releases the PrecivityAD2 blood test for clinical care (2023), (available at https://c2n.com/news-releases/cnnbspdiagnostics-releases-the-precivityad2-blood-test-for-clinical-care).

23. Roche Diagnostics. FDA cleared p tau181 Alzheimer’s blood test (2025), (available at https://diagnostics.roche.com/us/en/news-listing/2025/fda-cleared-ptau181-alzheimers-blood-test.html).

24. Roche receives FDA clearance for Elecsys® pTau217, advancing Alzheimer’s disease assessment across primary and specialty care. Accessed September 5, 2026, https://diagnostics.roche.com/us/en/news-listing/2026/fda-elecsys-ptau217-alzheimers-blood-test.html.

25. X. Cui, Z. Lu, The clinical translation of plasma p-tau217 warrants higher urgency than additional efficacy validation studies. Alzheimer’s & Dementia 21, e70477 (2025).

26. M. Olvera-Rojas et al., Influence of medical conditions on the diagnostic accuracy of plasma p-tau217 and p-tau217/Aβ42. Alzheimer’s & Dementia 21, e14430 (2025).

27. J. Yun et al., Plasma Phosphorylated Tau 217 Cutoffs for Amyloid Pathology and Kidney Function, Body Mass Index, and Anemia. JAMA Neurol, (2026).

28. M. M. Mielke et al., Performance of plasma phosphorylated tau 181 and 217 in the community. Nature Medicine 28, 1398–1405 (2022).

29. S. Abu-Rumeileh et al., Phosphorylated tau 181 and 217 are elevated in serum and muscle of patients with amyotrophic lateral sclerosis. Nature Communications 16, 2019 (2025).

30. F. Gonzalez-Ortiz et al., Plasma phosphorylated-tau217 is increased in Niemann–Pick disease type C. Brain Communications 6, (2024).

31. J. A. Bornhorst et al., Quantitative Assessment of the Effect of Chronic Kidney Disease on Plasma P-Tau217 Concentrations. Neurology 104, e210287 (2025).

32. T. Pushkarsky et al., Abundance of Nef and p-Tau217 in Brains of Individuals Diagnosed with HIV-Associated Neurocognitive Disorders Correlate with Disease Severance. Mol Neurobiol 59, 1088–1097 (2022).

33. S. A. Kaeser et al., Blood phosphorylated tau elevation as a biomarker in immunoglobulin light chain and transthyretin amyloidosis. Nature Medicine 32, 1676–1681 (2026).

34. H. Wesseling et al., Tau PTM Profiles Identify Patient Heterogeneity and Stages of Alzheimer&#x2019;s Disease. Cell 183, 1699–1713.e1613 (2020).

35. Y. Hirota et al., Biomarker-related phospho-tau217 appears in synapses around A&#x3b2; plaques prior to tau tangle in cerebral cortex of preclinical Alzheimer&#x2019;s disease. Cell Reports 44, (2025).

36. I. Fischer, Evolutionary perspective of Big tau structure: 4a exon variants of MAPT. Frontiers in Molecular Neuroscience Volume 15 **-** 2022, (2022).

37. A. Corsi, C. Bombieri, M. T. Valenti, M. G. Romanelli, Tau Isoforms: Gaining Insight into MAPT Alternative Splicing. International Journal of Molecular Sciences 23, 15383 (2022).

38. R. K. Koppisetti et al., Distribution of Big Tau Isoforms in the Human Central and Peripheral Nervous System. Annals of Neurology n/a.

39. I. Fischer, P. W. Baas, Big Tau: Structure, Evolutionary Divergence, and Emerging Roles in Cytoskeletal Dynamics and Tauopathies. Cells 15, 241 (2026).

40. I. S. Georgieff et al., Expression of high molecular weight tau in the central and peripheral nervous systems. Journal of Cell Science 105, 729–737 (1993).

41. B. N. Dugger et al., The Presence of Select Tau Species in Human Peripheral Tissues and Their Relation to Alzheimer’s Disease. Journal of Alzheimer’s Disease 51, 345–356 (2016).

42. M. Luciani et al., Big tau aggregation disrupts microtubule tyrosination and causes myocardial diastolic dysfunction: from discovery to therapy. European Heart Journal 44, 1560–1570 (2023).

43. W. Feng et al., NULISA: a proteomic liquid biopsy platform with attomolar sensitivity and high multiplexing. Nature Communications 14, 7238 (2023).

44. F. Gonzalez-Ortiz et al., Brain-derived tau: a novel blood-based biomarker for Alzheimer’s disease-type neurodegeneration. Brain 146, 1152–1165 (2022).

45. F. Gonzalez-Ortiz et al., Plasma brain-derived tau is an amyloid-associated neurodegeneration biomarker in Alzheimer’s disease. Nature Communications 15, 2908 (2024).

46. N. R. Barthélemy et al., Highly accurate blood test for Alzheimer’s disease is similar or superior to clinical cerebrospinal fluid tests. Nature Medicine 30, 1085–1095 (2024).

47. Y. Jiang et al., Head-to-head comparison of brain-derived pTau217 and total pTau217 for brain amyloid and tau pathology classification. Proceedings of the National Academy of Sciences 123, e2536792123 (2026).

48. J. R. Chong et al., Plasma brain-derived p-Tau217 outperforms other p-Tau species in detecting abnormal brain amyloid in an Asian cohort of older people with cerebrovascular disease burden. The Journal of Prevention of Alzheimer’s Disease 13, 100615 (2026).

49. R. A. Rissman et al., Plasma Aβ42/Aβ40 and phospho-tau217 concentration ratios increase the accuracy of amyloid PET classification in preclinical Alzheimer’s disease. Alzheimer’s & Dementia 20, 1214–1224 (2024).

50. M. R. Meyer et al., Clinical validation of the PrecivityAD2 blood test: A mass spectrometry-based test with algorithm combining %p-tau217 and Aβ42/40 ratio to identify presence of brain amyloid. Alzheimers Dement 20, 3179–3192 (2024).

51. C. Zoccali et al., Biomarkers in clinical epidemiology studies. Clinical Kidney Journal 17, (2024).

52. S. Palmqvist et al., Prediction of future Alzheimer’s disease dementia using plasma phospho-tau combined with other accessible measures. Nature Medicine 27, 1034–1042 (2021).

53. R. Ossenkoppele et al., Plasma p-tau217 and tau-PET predict future cognitive decline among cognitively unimpaired individuals: implications for clinical trials. Nat Aging 5, 883– 896 (2025).

54. R. A. Deek et al., Head-to-head comparison of plasma p-tau217 immunoassays for incipient Alzheimer’s disease in community cohorts. Alzheimer’s Research & Therapy 18, 61 (2026).

55. N. Warmenhoven et al., A comprehensive head-to-head comparison of key plasma phosphorylated tau 217 biomarker tests. Brain 148, 416–431 (2025).

56. N. J. Ashton et al., The Alzheimer’s Association Global Biomarker Standardization Consortium (GBSC) plasma phospho-tau Round Robin study. Alzheimer’s & Dementia 21, e14508 (2025).

57. R. La Joie et al., Treatment-related amyloid clearance (TRAC): a framework to characterize patients in the era of anti-amyloid therapies. Alzheimer’s & Dementia 21, e70997 (2025).

58. S. Vigneswaran et al., “Real-world” eligibility for anti-amyloid treatment in a tertiary memory clinic setting. Alzheimer’s & Dementia 21, e70375 (2025).

59. L. Martín-Aguilar et al., Big tau and brain-derived tau reveal peripheral and central nervous system involvement in neuropathies. medRxiv, 2026.2008.2027.26361202 (2026).

60. A. Biosciences. (2026).

61. M. N. Nafash et al., Plasma brain-derived tau: analytical and clinical validation of the first commercial immunoassay. Scientific Reports 16, 10124 (2026).

62. R. C. Petersen et al., Alzheimer’s Disease Neuroimaging Initiative (ADNI). Neurology 74, 201–209 (2010).

63. M. W. Weiner et al., The Alzheimer’s Disease Neuroimaging Initiative: Progress report and future plans. Alzheimer’s & Dementia 6, 202–211.e207 (2010).

64. O. L. Lopez et al., Research evaluation and diagnosis of probable Alzheimer’s disease over the last two decades: I. Neurology 55, 1854–1862 (2000).

65. J. C. Morris et al., The Uniform Data Set (UDS): clinical and cognitive variables and descriptive data from Alzheimer Disease Centers. Alzheimer Dis Assoc Disord 20, 210–216 (2006).

66. Consensus recommendations for the postmortem diagnosis of Alzheimer’s disease. The National Institute on Aging, and Reagan Institute Working Group on Diagnostic Criteria for the Neuropathological Assessment of Alzheimer’s Disease. Neurobiol Aging 18, S1–2 (1997).

67. S. S. Mirra et al., The Consortium to Establish a Registry for Alzheimer’s Disease (CERAD). Neurology 41, 479–479 (1991).

68. H. Braak, E. Braak, Neuropathological stageing of Alzheimer-related changes. Acta Neuropathol 82, 239–259 (1991).

69. A. D. Cohen et al., Connectomics in Brain Aging and Dementia - The Background and Design of a Study of a Connectome Related to Human Disease. Front Aging Neurosci 13, 669490 (2021).

70. W. E. Klunk et al., Imaging brain amyloid in Alzheimer’s disease with Pittsburgh Compound-B. Ann Neurol 55, 306–319 (2004).

71. B. J. Lopresti et al., Comparison of visual and analytic methods of Alzheimer’s disease pathologic staging. Alzheimer’s & Dementia 21, e70520 (2025).

72. P. Bourgeat et al., Implementing the centiloid transformation for (11)C-PiB and β-amyloid (18)F-PET tracers using CapAIBL. Neuroimage 183, 387–393 (2018).

73. W. J. Jagust et al., The ADNI PET Core at 20. Alzheimers Dement 20, 7340–7349 (2024).

74. P.-F. Meyer et al., Characterization of Alzheimer Disease Biomarker Discrepancies Using Cerebrospinal Fluid Phosphorylated Tau and AV1451 Positron Emission Tomography. JAMA Neurology 77, 508–516 (2020).

75. M. I. Kamboh et al., Population-based genome-wide association study of cognitive decline in older adults free of dementia: identification of a novel locus for the attention domain. Neurobiol Aging 84, 239.e215–239.e224 (2019).

76. Y. Benjamini, Y. Hochberg, Controlling the False Discovery Rate: A Practical and Powerful Approach to Multiple Testing. Journal of the Royal Statistical Society: Series B (Methodological) 57, 289–300 (2018).

77. E. R. DeLong, D. M. DeLong, D. L. Clarke-Pearson, Comparing the areas under two or more correlated receiver operating characteristic curves: a nonparametric approach. Biometrics 44, 837–845 (1988).

