## Supplementary Material for "CNS-selective plasma p-tau217 accurately captures Alzheimer’s disease pathology and progression"

Zeng X *et al*.

**The PDF file includes:**

Figs. S1 to S8

Table S1 to S8

**Other Supplementary Material for this manuscript includes the following:**

Data S1 to S6

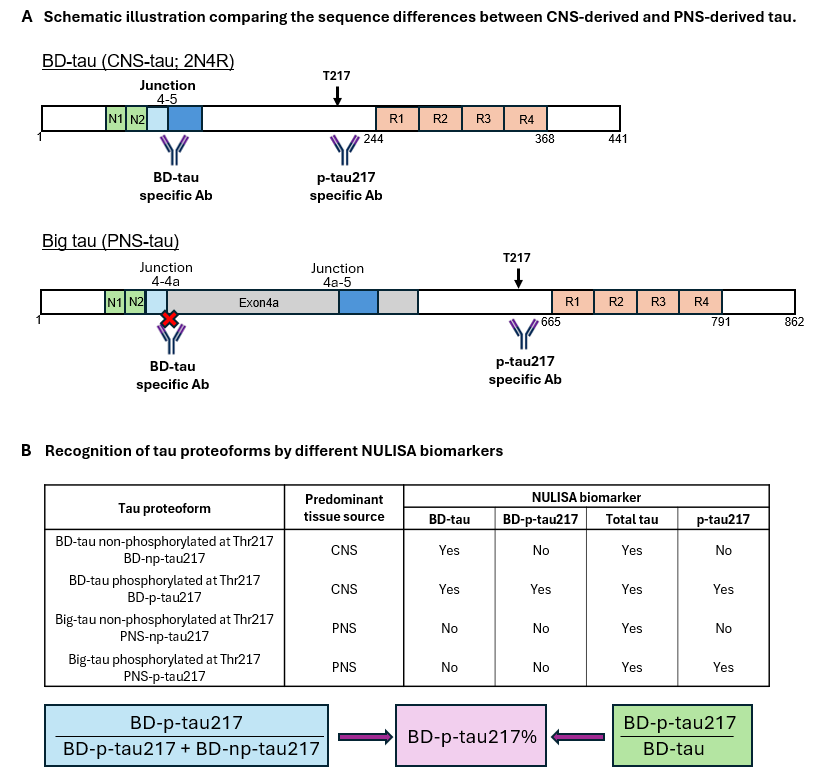

**Fig. S1. Conceptual basis of the BD‑p‑tau217% assay.** **(A)** Schematic illustration comparing sequence features of CNS‑derived tau (BD-tau) and PNS‑derived tau (Big-tau), highlighting regions relevant for BD‑tau antibody recognition and the phosphorylation site at Thr217. The BD-p-tau217 immunoassay combines two antibodies, one that is specific for p-tau217 and another that binds specifically at the exon4-5 junction. **(B)** Table summarizing the recognition of tau proteoforms by different NULISA tau biomarkers, including BD‑tau BD‑p‑tau217, total-tau, and p‑tau217. Proportion of BD-tau phosphorylated at Thr217, abbreviated as BD-p-tau217%, was quantified by the ratio of two NULISA biomarkers, BD-p-tau217/BD-tau. Note that T217 refers to the threonine position based on the tau 2N4R isoform. The corresponding residue in big‑tau (GENCODE transcript ENST00000262410.10) is Thr609. In the NULISA panel, BD‑tau and total tau are labeled as BD‑MAPT and MAPT, respectively. “np” denotes the non‑phosphorylated form.

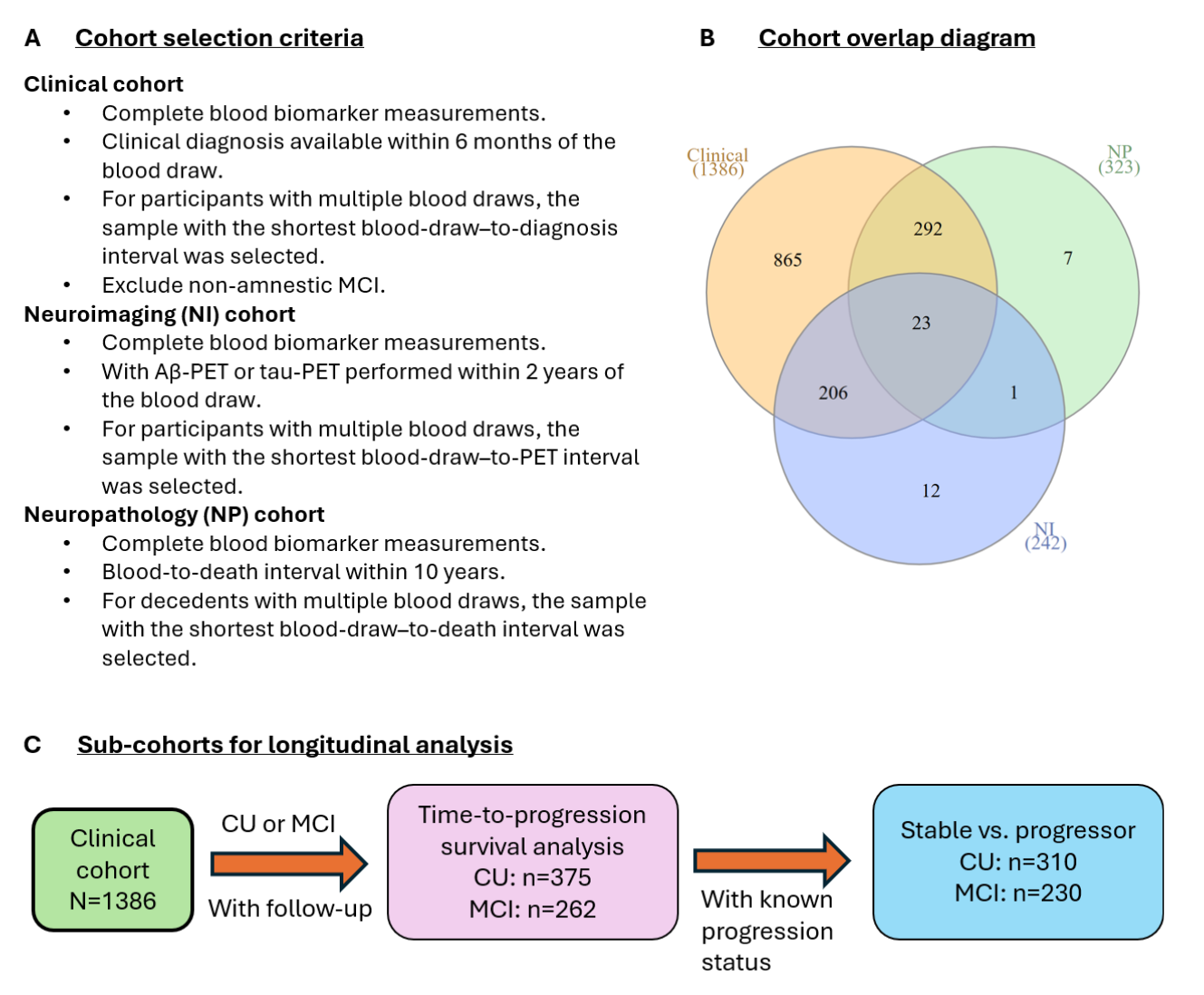
**Fig. S2. Participant selection, cohort overlap, and derivation of longitudinal analysis sub‑cohorts in the Pitt ADRC.** All participants were drawn from the Pitt‑ADRC study. Panel A outlines the participant selection criteria for the three cohorts included in this study (clinical, neuropathological, and neuroimaging). Panel B presents the Venn diagram illustrating the overlap of participants across these cohorts. Panel C depicts the selection of participants for longitudinal analyses. Progression status was defined based on clinical change within a 5‑year window. Cognitively unimpaired (CU) participants at baseline who progressed to mild cognitive impairment (MCI) or Alzheimer’s dementia (AD) within 5 years were classified as progressors. Participants with baseline MCI who progressed to AD within 5 years were likewise classified as progressors. Individuals whose clinical status remained unchanged at their first follow‑up visit beyond 5 years after baseline were classified as stable.

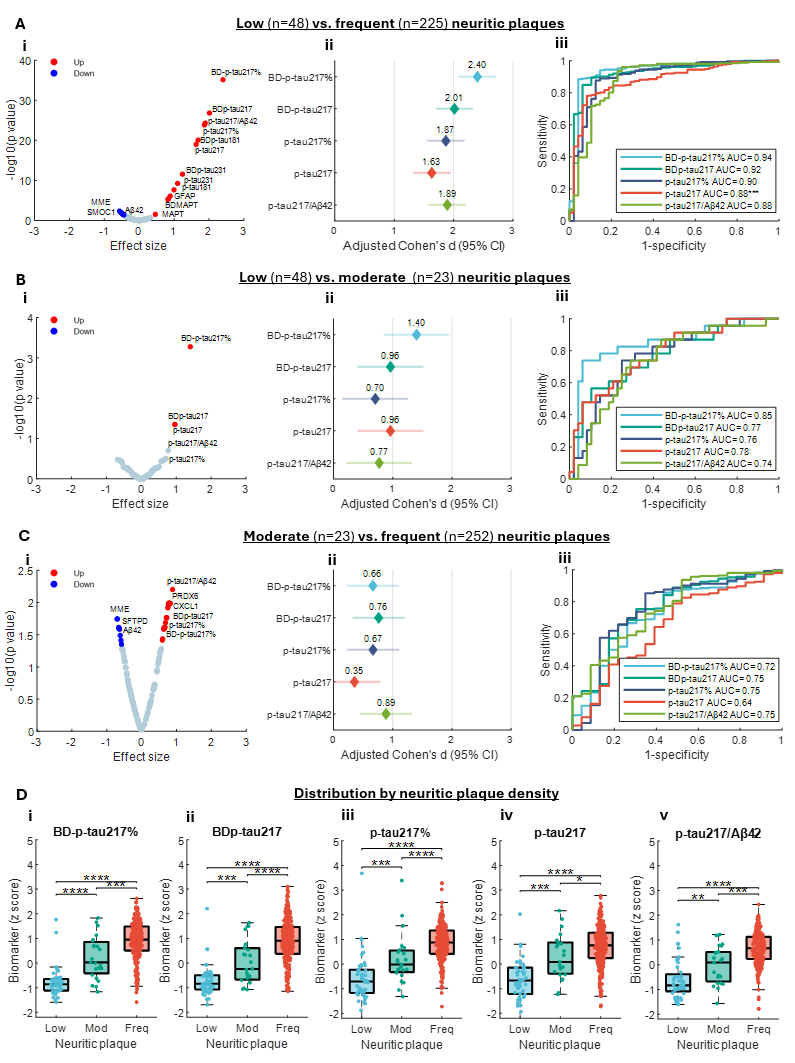

**Figure S3. Association of biomarker levels with neuritic plaque density in the Pitt-ADRC neuropathology cohort.** Panels A–C compare biomarker levels between low vs. frequent, low vs. moderate, and intermediate vs. frequent neuritic plaque density, respectively. Volcano plots display age‑ and sex‑adjusted Cohen’s *d* effect sizes on the x‑axis and -log₁₀(p values) on the y‑axis. Red dots indicate up‑regulated and blue dots downregulated in participants with higher neuritic plaque density. P values were FDR-adjusted using Benjamini-Hochberg procedure. Forest plots depict the point estimates and 95% confidence intervals of Cohen’s *d*. Biomarker levels were z-normalized. AUC plots were based on logistic regression classification performance using biomarker level alone. Asterisks denote DeLong test comparisons with BD-p‑tau217% as the reference biomarker, with *, **, and *** indicating p < 0.05, 0.01, and 0.001, respectively. Panels D show boxplot distributions of biomarker levels across neuritic plaque categories; the center line indicates the median, box edges represent the 25th and 75th percentiles, and whiskers extend to the most extreme non‑outlier values. Asterisks denote significance levels for pairwise rank‑sum tests after Benjamini–Hochberg correction, with *, **, ***, and **** for p < 0.05, 0.01, 0.001, and 0.0001, respectively.

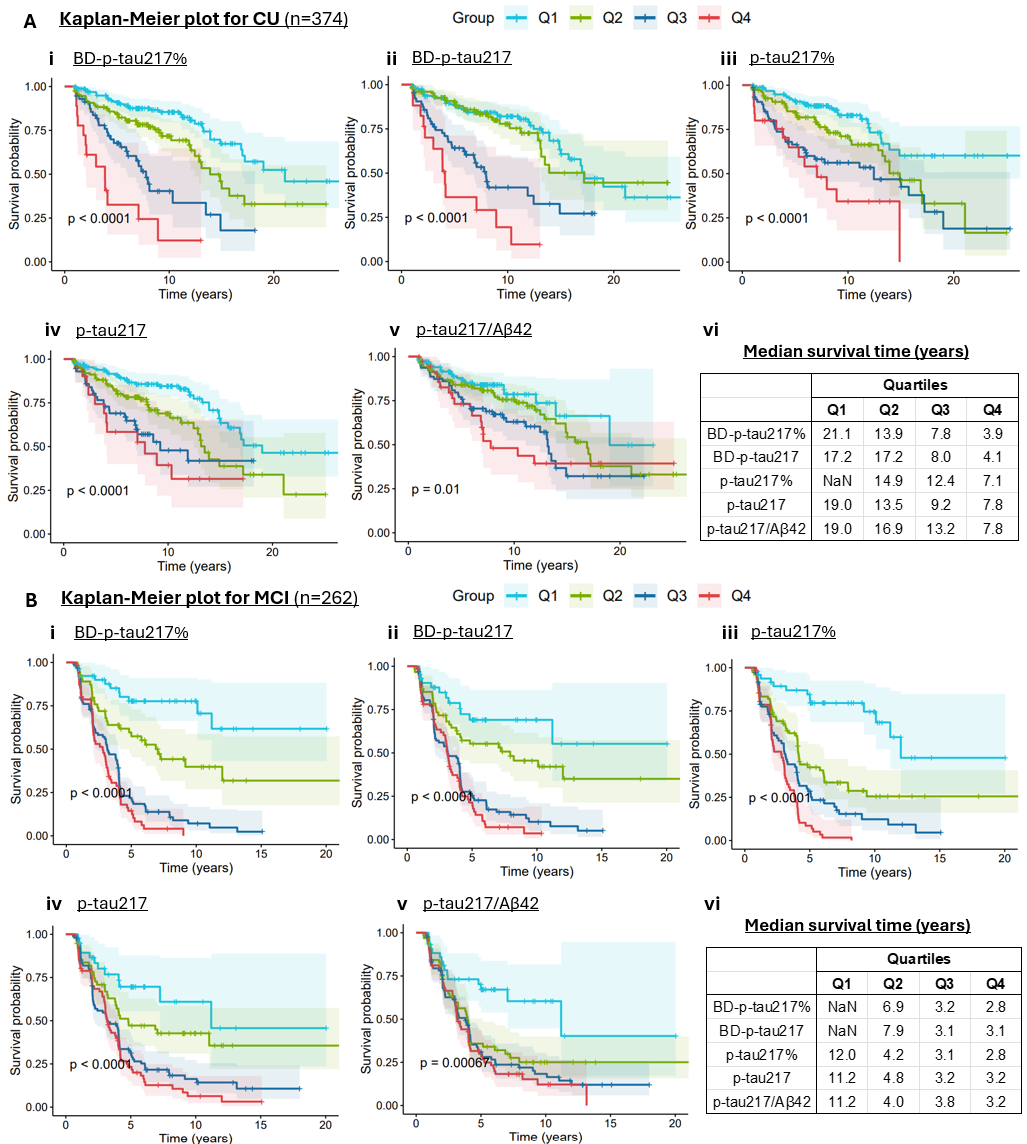

**Fig. S4. Performance of biomarkers in predicting future clinical progression in the Pitt-ADRC clinical cohort.** Shown are Kaplan-Meier survival curves illustrating time‑to‑incident clinical progression for individuals who were CU (**A**) or had MCI (**B**) at baseline, stratified by biomarker quartiles in the full cohort. CU, clinically unimpaired; MCI, mild cognitive impairment. Shaded area indicate 95% confidence interval.

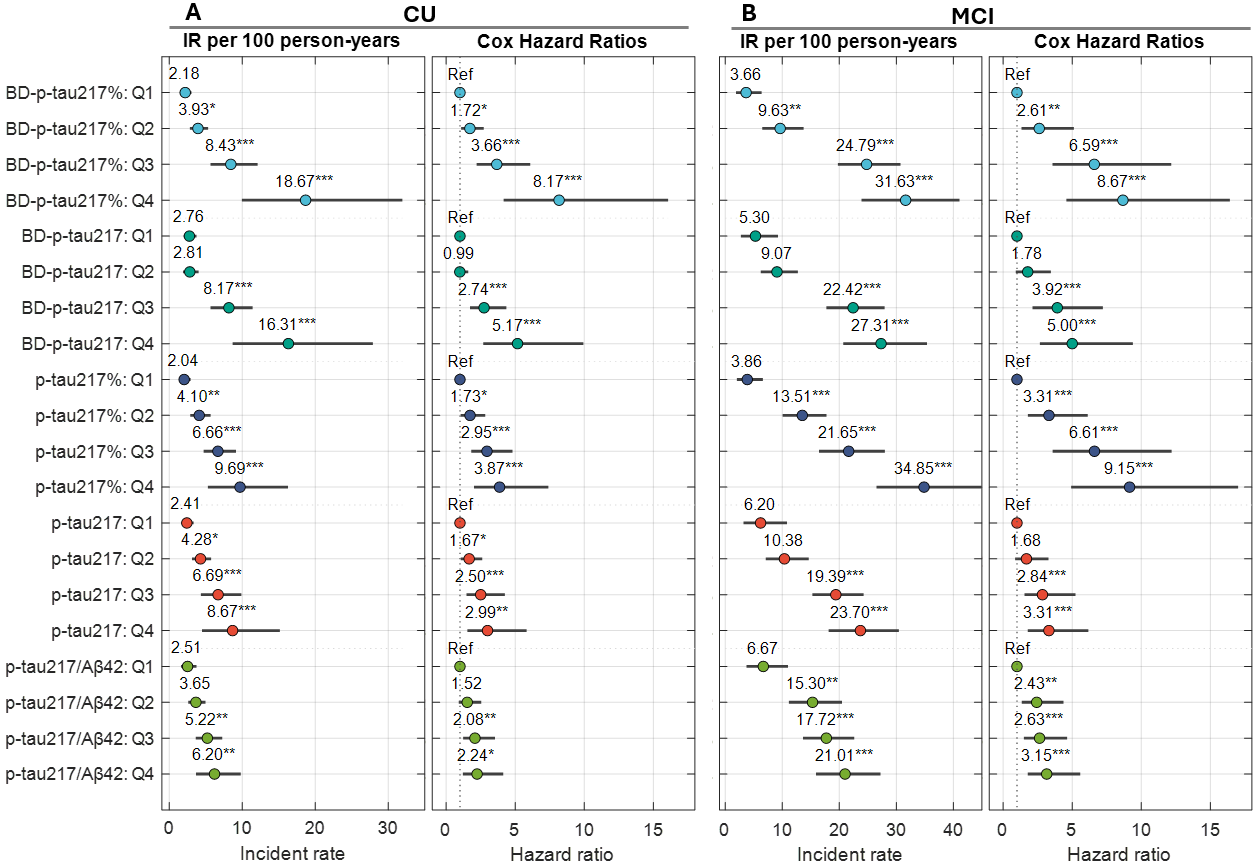

**Fig. S5. Forest plots of incidence rate (IR) and hazard ratio (HR) in predicting future clinical progression in the Pitt-ADRC clinical cohort.** HRs were estimated using Cox proportional hazards models adjusted for age and sex, with quartile-1 (Q1) as the reference group. Asterisks indicate significance relative to Q1, with *, **, and *** corresponding to p < 0.05, 0.01, and 0.001, respectively. Circles represent point estimates, and horizontal lines represent 95% confidence intervals.

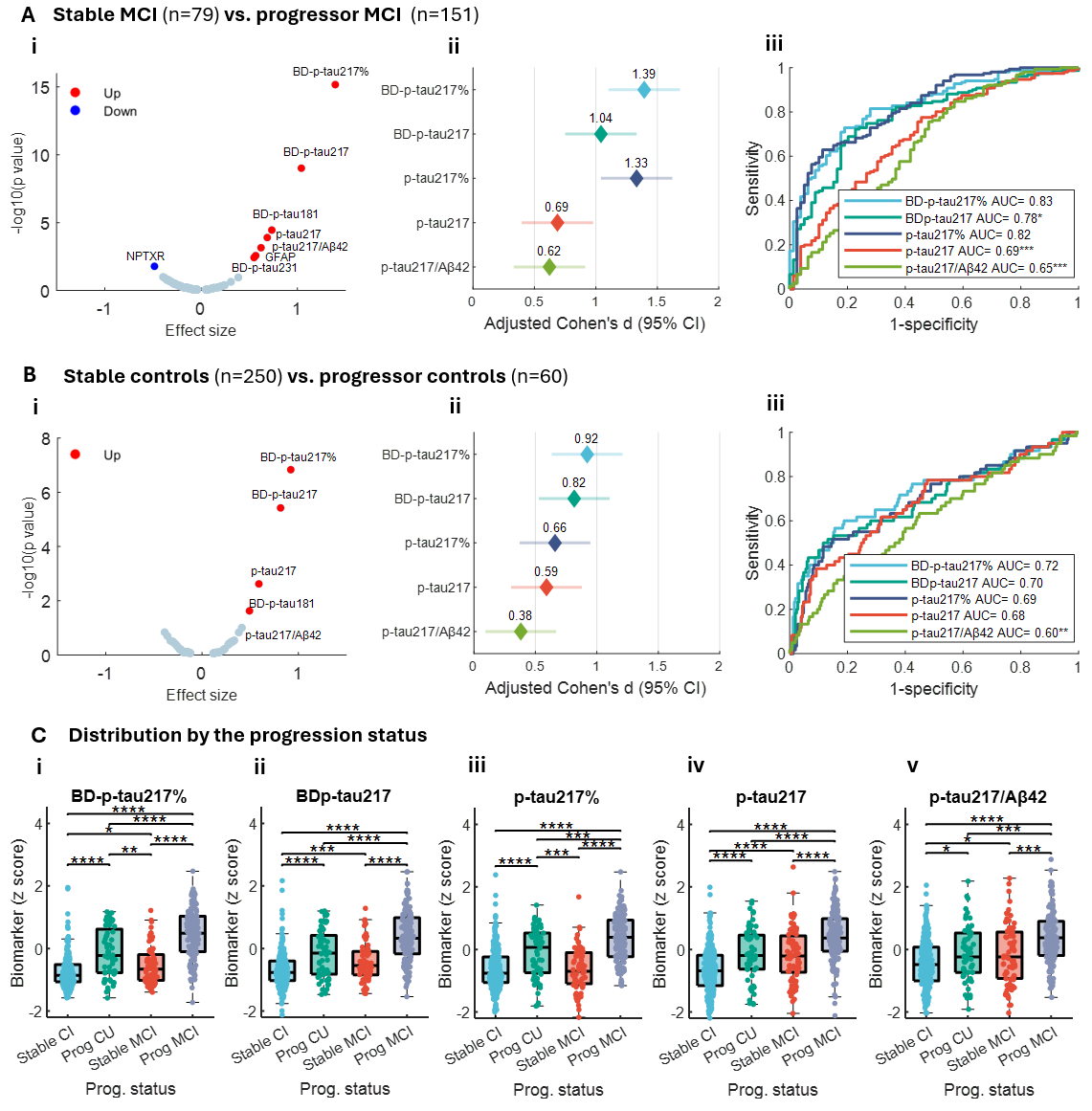

**Fig. S6. Association of biomarker levels with clinical progression status in the Pitt-ADRC clinical cohort.** Panels A-B compare biomarker levels between stable MCI vs. progressor MCI, stable controls vs. progressor controls, respectively. Volcano plots display age‑ and sex‑adjusted Cohen’s d effect sizes on the x‑axis and -log₁₀(p values) on the y‑axis. Red dots indicate up-regulated biomarkers and blue dots indicate those downregulated in progressors. P values were FDR-adjusted using Benjamini-Hochberg procedure. Forest plots depict the point estimates, and 95% confidence intervals of Cohen’s d. Biomarker levels were z-normalized. AUC plots were based on logistic regression classification performance using biomarker level alone. Asterisks denote DeLong test comparisons with BD-p‑tau217% as the reference biomarker, with *, **, and *** indicating p < 0.05, 0.01, and 0.001, respectively. Panels C show boxplot distributions of biomarker levels by progression status; the center line indicates the median, box edges represent the 25th and 75th percentiles, and whiskers extend to the most extreme non‑outlier values. Asterisks denote significance levels for pairwise rank‑sum tests after Benjamini–Hochberg correction, with *, **, ***, and **** for p < 0.05, 0.01, 0.001, and 0.0001, respectively.

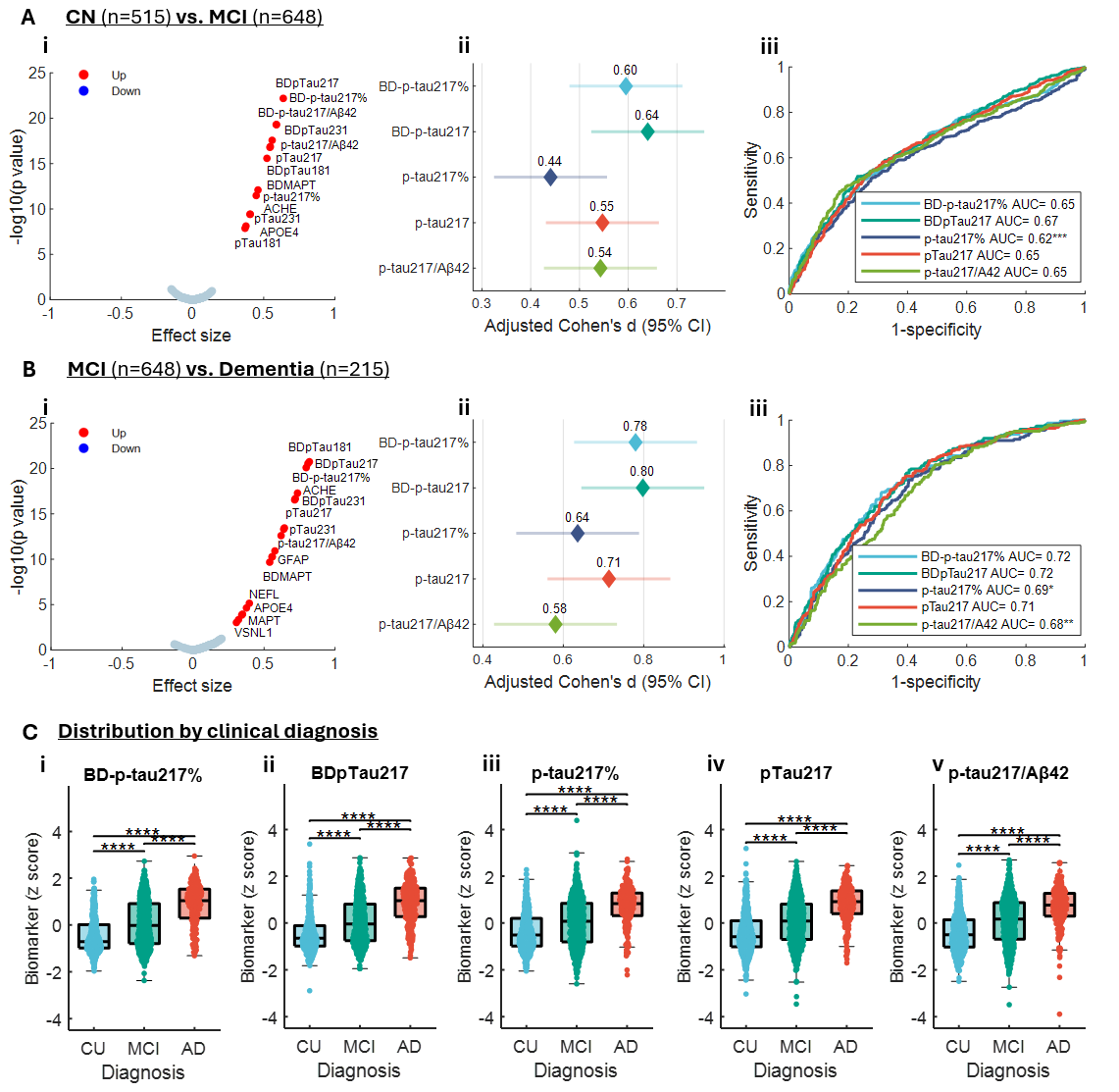

**Fig. S7. Association of biomarker levels with clinical progression status in the ADNI cohort.** Panels A-C compare biomarker levels between cognitive unimpaired (CU) vs. Alzheimer disease (AD), CU vs. mild cognitive impairment (MCI), and MCI vs. AD, respectively. Volcano plots display age‑ and sex‑adjusted Cohen’s d effect sizes on the x‑axis and -log₁₀(p values) on the y‑axis. Red dots indicate up‑regulated, and blue dots indicate downregulated. P values were FDR-adjusted using Benjamini-Hochberg procedure. Forest plots depict the point estimates, and 95% confidence intervals of Cohen’s d. Biomarker levels were z-normalized. AUC plots were based on logistic regression classification performance using biomarker level alone. Asterisks denote DeLong test comparisons with BD-p‑tau217% as the reference biomarker, with *, **, and *** indicating p < 0.05, 0.01, and 0.001, respectively. Panels Di-v show boxplot distributions of biomarker levels by diagnosis status; the center line indicates the median, box edges represent the 25th and 75th percentiles, and whiskers extend to the most extreme non‑outlier values. Asterisks denote significance levels for pairwise rank‑sum tests after Benjamini–Hochberg correction, with *, **, ***, and **** for p < 0.05, 0.01, 0.001, and 0.0001, respectively.

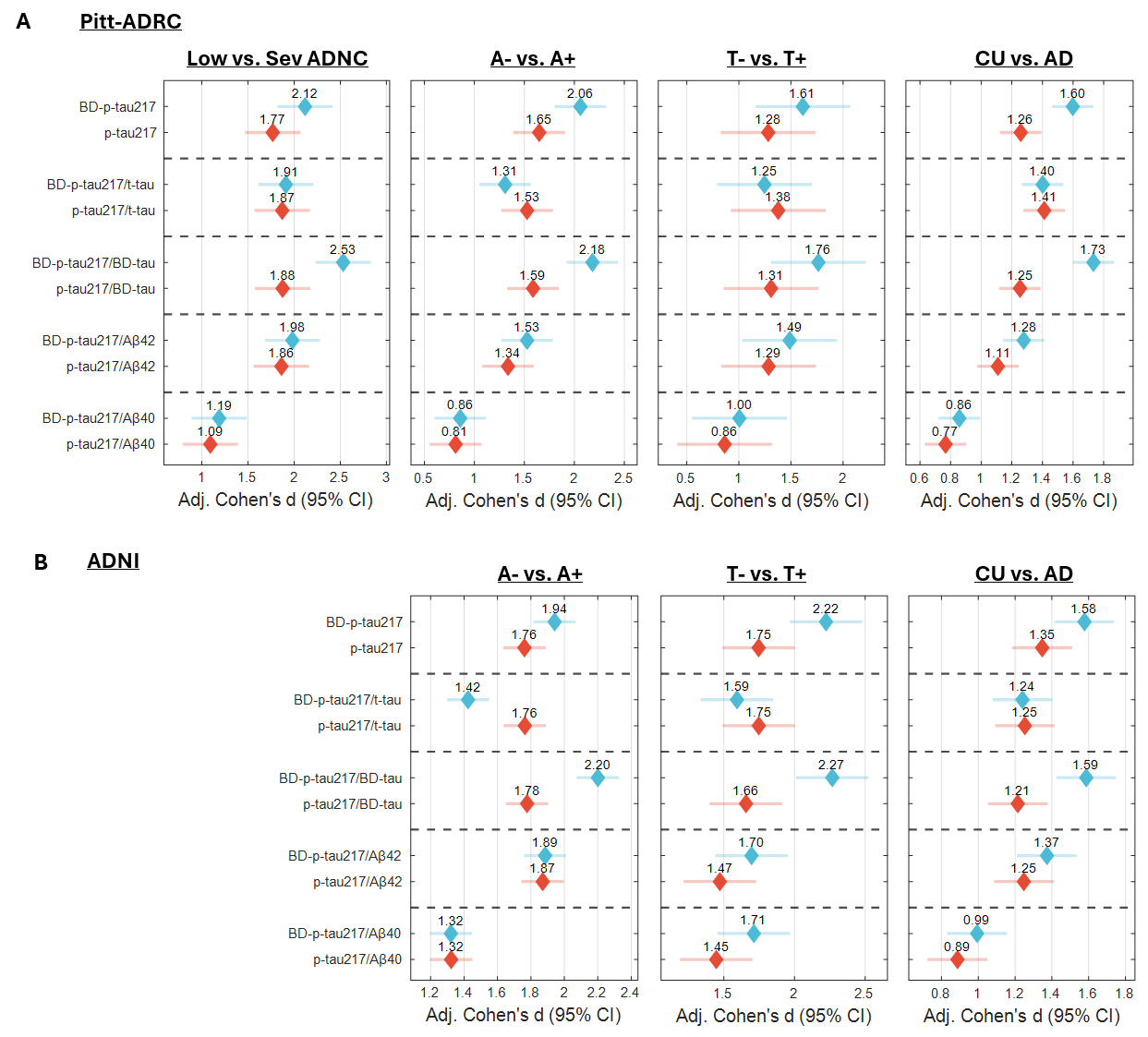

**Fig. S8. Comparison of alternative biomarker normalization strategies.** (A) Forest plots showing age- and sex-adjusted effect sizes for discriminating low versus severe ADNC, A− versus A+, T− versus T+, and CU versus AD participants in the Pitt-ADRC cohort. (B) Forest plots showing age- and sex-adjusted effect sizes for discriminating A− versus A+, T− versus T+, and CU versus AD participants in the ADNI cohort. Forest plots display adjusted Cohen’s d estimates and 95% confidence intervals. Biomarker concentrations were z-normalized prior to effect size calculation.

**Table S1. Participant characteristics for the Pitt-ADRC neuropathology cohort stratified by ADNC.**

|  | **Total** | **Low** | **Intermediate** | **High** | **p-value** |
| --- | --- | --- | --- | --- | --- |
| Number of participants, n | 323 | 56 | 47 | 220 |  |
| Age (blood; years) | 74.5 (66.0-80.6) | 68.7 (62.9-78.3) | 80.6 (76.9-83.6) | 74.0 (65.5-79.7) | 2.42E-07 |
| Age (death; years) | 79.7 (70.0-85.0) | 73.0 (67.5-83.0) | 85.0 (80.3-89.8) | 79.0 (69.0-85.0) | 5.57E-07 |
| Blood-to-death (years) | 5.3 (3.5-7.1) | 4.5 (2.9-6.5) | 4.8 (3.5-6.8) | 5.6 (3.6-7.1) | 0.060 |
| Sex |  |  |  |  | 1.54E-04 |
| Female | 158 (48.9%) | 19 (33.9%) | 19 (40.4%) | 120 (54.6%) |  |
| Male | 165 (51.1%) | 37 (66.1%) | 28 (59.6%) | 100 (45.5%) |  |
| Education (years) | 15 (12-17) | 16 (12-18) | 16 (12-18) | 15 (12-16) | 0.430 |
| Race |  |  |  |  | 0.085 |
| NHW | 316 (97.8%) | 54 (96.4%) | 46 (97.9%) | 216 (98.2%) |  |
| B/AA | 6 (1.9%) | 2 (3.6%) | 1 (2.1%) | 3 (1.4%) |  |
| Others/missing | 1 (0.3%) | 0 | 0 | 1 (0.5%) |  |
| MMSE | 21 (16-24) | 23 (17.5-26.0) | 23 (17.3-26.0) | 20 (15-23) | 9.84E-05 |
| Clinical diagnosis^a^ |  |  |  |  | 1.14E-25 |
| CU | 12 (3.8%) | 3 (5.7%) | 5 (11.1%) | 4 (1.8%) |  |
| MCI | 21 (6.67%) | 3 (5.7%) | 5 (11.1%) | 13 (6.0%) |  |
| AD dementia | 238 (75.6%) | 15 (28.3%) | 31 (68.9%) | 192 (88.5%) |  |
| Non-AD dementia | 44 (14.0) | 32 (60.4%) | 4 (8.9%) | 8 (3.7%) |  |
| Missing | 8 | 3 | 2 | 3 |  |
| CDR-GS^a^ |  |  |  |  | 0.232 |
| 0 | 12 (3.8%) | 4 (7.4%) | 4 (8.5%) | 4 (1.8%) |  |
| 0.5 | 78 (24.5%) | 14 (25.9%) | 13 (27.7%) | 51 (23.4%) |  |
| 1 | 150 (47.0%) | 27 (50.0%) | 19 (40.4%) | 104 (47.7%) |  |
| 2 | 64 (20.1%) | 7 (13.0%) | 10 (21.3%) | 47 (21.6%) |  |
| 3 | 15 (4.7%) | 2 (3.7%) | 1 (2.1%) | 12 (5.5%) |  |
| Missing | 4 | 2 | 0 | 2 |  |
| *APOE4* carrier |  |  |  |  | 2.04E-06 |
| No | 149 (46.3%) | 41 (74.6%) | 24 (51.1%) | 84 (38.2%) |  |
| Yes | 173 (53.7%) | 14 (25.5%) | 23 (48.9%) | 136 (61.8%) |  |
| Missing | 1 | 1 | 0 | 0 |  |
| NFT Braak |  |  |  |  | 1.55E-131 |
| Low | 34 (10.9%) | 34 (75.6%) | 0 | 0 |  |
| PART | 11 (3.5%) | 11 (24.4%) | 0 | 0 |  |
| Middle | 47 (15.1%) | 0 | 47 (100.0%) | 0 |  |
| High | 220 (70.5%) | 0 | 0 | 220 (100.0%) |  |
| Missing | 11 | 11 | 0 | 0 |  |
| CERAD rating |  |  |  |  | 5.18E-70 |
| Low | 48 (14.9%) | 48 (85.7%) | 0 | 0 |  |
| Moderate | 23 (7.1%) | 2 (3.6%) | 16 (34.0%) | 5 (2.3%) |  |
| Frequent | 252 (78.0%) | 6 (10.7%) | 31 (66.0%) | 215 (97.7%) |  |

NHW, Non-Hispanic White; B/AA, Black/African-American; CU, clinically unimpaired; AD, Alzheimer’s disease; MCI, mild cognitive impairment; PET, positron emission tomography; MMSE, Mini–Mental State Examination; CDR-GS, Clinical Dementia Rating global score; ADNC, Alzheimer’s disease neuropathologic change; NFT, neurofibrillary tangle; PART, primary age‑related tauopathy. Categorical variables are presented as count (percentage), and continuous variables as median and interquartile range. P‑values were calculated using the Kruskal–Wallis test for continuous variables and Fisher’s exact test or the chi‑square test for categorical variables. ^a^ Status based on the nearest visit within 6 months of blood draw.

**Table S2. Participant characteristics for the Pitt-ADRC neuroimaging cohort by Aβ-PET status.**

|  | **Total** | **Aβ-PET-** | **Aβ-PET+** | **p-value** |
| --- | --- | --- | --- | --- |
| Number of participants, n | 242^b^ | 113 | 128 |  |
| Age (blood; years) | 68.5 (61.7-74.5) | 65.5 (60.0-71.8) | 70.3 (63.2-75.5) | 0.003 |
| Sex |  |  |  | 0.895 |
| Female | 104 (43.0%) | 48 (42.5%) | 56 (43.8%) |  |
| Male | 138 (57.0%) | 65 (57.5%) | 72 (56.3%) |  |
| Education (years) | 16 (14-18) | 16 (14-18) | 16 (14-18) | 0.772 |
| Race |  |  |  | 1^c^ |
| NHW | 232 (95.9%) | 109 (96.5%) | 122 (95.3%) |  |
| B/AA | 9 (3.7%) | 4 (3.5%) | 5 (3.9%) |  |
| Others/missing | 1 (0.4%) | 0 | 1 (0.8%) |  |
| MMSE | 27 (23-29) | 28 (26-29) | 25 (21-28) | 3.68E-07 |
| Clinical diagnosis^a^ |  |  |  | 6.09E-06 |
| CU | 64 (27.6%) | 46 (43.8%) | 18 (14.3%) |  |
| MCI | 70 (30.2%) | 34 (32.4%) | 36 (28.6%) |  |
| AD dementia | 72 (31.0%) | 9 (8.6%) | 63 (50.0%) |  |
| Non-AD dementia | 26 (11.2%) | 16 (15.2%) | 9 (7.1%) |  |
| missing | 10 | 8 | 2 |  |
| CDR-GS^a^ |  |  |  | 5.20E-06 |
| 0 | 39 (16.1%) | 23 (20.4%) | 16 (12.5%) |  |
| 0.5 | 151 (61.4%) | 77 (68.1%) | 73 (57.0%) |  |
| 1 | 42 (17.4%) | 8 (7.1%) | 34 (26.6%) |  |
| 2 | 10 (4.1%) | 5 (4.4%) | 5 (3.9%) |  |
| *APOE4* carrier |  |  |  | 3.08E-05 |
| No | 137 (59.0%) | 78 (73.6%) | 58 (46.4%) |  |
| Yes | 95 (41.0%) | 28 (26.4%) | 67 (53.6%) |  |
| Missing | 10 | 7 | 3 |  |
| PiB Centiloid | 25.7 (-1.34-77.57) | -1.54 (-7.26-2.84) | 75.25 (54.09-96.64) | 6.96E-41 |
| Tau PET |  |  |  | 6.93E-10 |
| Negative | 39 (45.9%) | 32 (80.0%) | 6 (13.6%) |  |
| Positive | 46 (54.1%) | 8 (20.0%) | 38 (86.4%) |  |
| Missing | 154 | 68 | 86 |  |
| Tau PET SUVR | 1.19 (1.11-1.57) | 1.12 (1.07-1.16) | 1.57 (1.23-1.93) | 3.95E-10 |

NHW, Non-Hispanic White; B/AA, Black/African-American; AD, Alzheimer’s disease; CU, cognitively unimpaired; MCI, mild cognitive impairment; PET, positron emission tomography; MMSE, Mini–Mental State Examination; CDR-GS, Clinical Dementia Rating global score; SUVR, standardized uptake value ratio. Categorical variables are presented as count (percentage), and continuous variables as median and interquartile range. P‑values were calculated using the Kruskal–Wallis test for continuous variables and Fisher’s exact test or the chi‑square test for categorical variables. ^a^ Status based on the nearest visit within 6 months of blood draw. ^b^ One sample had tau-PET but no Aβ-PET. ^c^ Only NHW and B/AA were included for p-value calculation.

**Table S3. Participant characteristics for the Pitt-ADRC clinical cohort stratified by clinical diagnosis.**

|  | **Total** | **CU** | **MCI** | **AD dementia** | **Non-AD dementia** | **p value** |
| --- | --- | --- | --- | --- | --- | --- |
| Number of participants, n | 1386 | 412 | 287 | 480 | 207 |  |
| Age (baseline; years) | 71.4 (64.7-77.5) | 69.2 (64.0-73.8) | 72.6 (67.6-78.2) | 75.0 (66.5-80.1) | 67.5 (61.0-75.0) | 5.52E-18 |
| Sex |  |  |  |  |  | 9.45E-06 |
| Female | 735 (53.0%) | 262 (63.6%) | 127 (44.3%) | 266 (55.4%) | 80 (38.7%) |  |
| Male | 651 (47.0%) | 150 (36.4%) | 160 (55.8%) | 214 (44.6%) | 127 (61.4%) |  |
| Education (years) | 16 (12-18) | 16 (14-18) | 16 (13-18) | 14 (12-16) | 14 (12-17) | 3.17E-15 |
| Race |  |  |  |  |  | 2.84E-06^b^ |
| NHW | 1268 (91.5%) | 344 (83.5%) | 275 (95.8%) | 450 (93.8%) | 199 (96.1%) |  |
| B/AA | 110 (7.9%) | 65 (15.8%) | 9 (3.1%) | 28 (5.8%) | 8 (3.9%) |  |
| Others/missing | 8 (0.6%) | 3 (0.7%) | 3 (1.1%) | 2 (0.4%) | 0 |  |
| MMSE | 26 (21-28) | 29 (28-30) | 27 (25-28) | 20 (16-23) | 22 (18-26) | 1.60E-176 |
| CDR-GS^a^ |  |  |  |  |  | 8.31E-05 |
| 0 | 308 (22.4%) | 296 (73.1%) | 8 (2.8%) | 1 (0.2%) | 3 (1.5%) |  |
| 0.5 | 570 (41.4%) | 108 (26.7%) | 269 (93.7%) | 132 (27.6%) | 61 (29.8%) |  |
| 1 | 345 (25.1%) | 1 (0.3%) | 10 (3.5%) | 241 (50.3%) | 93 (45.4%) |  |
| 2 | 130 (9.5%) | 0 | 0 | 89 (18.6%) | 41 (20.0%) |  |
| 3 | 23 (1.7%) | 0 | 0 | 16 (3.3%) | 7 (3.4%) |  |
| Missing | 10 | 7 | 0 | 1 | 2 |  |
| *APOE4* carrier |  |  |  |  |  | 4.00E-06 |
| No | 731 (53.4%) | 265 (65.1%) | 135 (47.5%) | 198 (41.8%) | 133 (64.9%) |  |
| Yes | 639 (46.6%) | 142 (34.9%) | 149 (52.5%) | 276 (58.2%) | 72 (35.1%) |  |
| Missing | 16 | 5 | 3 | 6 | 2 |  |
| Comorbidity, n(%)[missing]^c^ |  |  |  |  |  |  |
| Obesity^d^ | 250 (27.1%)[464] | 83 (31.8%) | 52(23.2%) | 73(23.2%) | 42(34.4%) | 0.015 |
| Diabetes | 163(17.2%)[439] | 52(18.3%) | 31(16.2%) | 55(17.1%) | 25(16.8%) | 0.947 |
| CVD | 205(21.7%)[439] | 72(25.3%) | 33(17.3%) | 60(18.6%) | 40(26.9%) | 0.038 |
| Hypertension | 452(49.2%)[467] | 140(50.7%) | 84(44.7%) | 144(45.9%) | 84(59.6%) | 0.026 |
| Hypercholesterolemia | 562(60.6%)[458] | 174(62.1%) | 105(55.9%) | 189(60.0%) | 94(64.8%) | 0.364 |
| Active depression | 289(30.6%)[442] | 82(28.8%) | 60(31.6%) | 101(31.5%) | 46(31.1%) | 0.882 |

NHW, Non-Hispanic White; B/AA, Black/African-American; AD, Alzheimer’s disease; CU, clinically unimpaired; MCI, mild cognitive impairment; MMSE, Mini–Mental State Examination; CDR, Clinical Dementia Rating. Categorical variables are presented as count (percentage), and continuous variables as median and interquartile range. P‑values were calculated using the Kruskal–Wallis test for continuous variables and Fisher’s exact test or the chi‑square test for categorical variables. Obesity was defined as a body mass index (BMI) ≥ 30. All percentages were calculated based on participants with non‑missing data for the corresponding variable. ^a^ Status based on the nearest visit within 6 months of blood draw. ^b^ Only NHW and B/AA were included for p-value calculation. ^c^ For comorbidities, n (%) reflects the number of participants with the specified comorbidity and the corresponding proportion among those with available data for that condition. The number shown in brackets [] indicates the number of participants with missing information for the respective comorbidity. ^d^ Obesity was defined as a body mass index (BMI) ≥ 30.

**Table S4. Participant characteristics for the ADNI cohort.**

| **Characteristics** | **Full cohort** | **Aβ-PET** | **Tau-PET** |
| --- | --- | --- | --- |
| Number of participants, n | 1428 | 999 | 360 |
| Age (blood; years), median (IQR) | 72.8 (10.3) | 71.5 (9.8) | 70.0 (9.0) |
| Female, n (%) | 676 (47.3%) | 505 (50.6%) | 197 (54.7%) |
| White, n (%) | 1356 (95.3%) | 949 (95.5%) | 329 (92.7%) |
| Education (years), median (IQR) | 16 (4) | 16 (4) | 16 (3) |
| *APOE4* carriers, n (%) | 648 (45.4%) | 440 (44.0%) | 153 (42.5%) |
| MMSE (baseline), median (IQR) | 28 (4) | 29 (3) | 29 (3) |
| Clinical diagnosis, n (%) |  |  |  |
| CU | 529 (37.0%) | 405 (40.5%) | 201 (55.8%) |
| MCI | 676 (47.3%) | 474 (47.5%) | 121 (33.6%) |
| Dementia | 223 (15.6%) | 120 (12.0%) | 38 (10.6%) |
| CDR-GS, n (%) |  |  |  |
| 0 | 526 (37.0%) | 405 (40.5%) | 202 (56.1%) |
| 0.5 | 784 (55.1%) | 527 (52.8%) | 138 (38.3%) |
| 1 | 111 (7.8%) | 66 (6.6%) | 20 (5.6%) |
| 2 | 1 (0.1%) | 1 (0.1%) | 0 |
| Aβ-PET (Neg/Pos), n | 504/495 | 504/495 | 201/154 |
| Tau-PET (Neg/Pos), n | 286/74 | 282/73 | 286/74 |

B/AA, Black/African-American; CU, cognitively unimpaired; MCI, mild cognitive impairment; PET, positron emission tomography; MMSE, Mini–Mental State Examination; CDR-GS, Clinical Dementia Rating global score.

**Table S5. Participant characteristics for the ADNI cohort stratified by Aβ-PET status.**

|  | **Total** | **Aβ-PET-** | **Aβ-PET+** | **p-value** |
| --- | --- | --- | --- | --- |
| Number of participants, n | 999 | 504 | 495 |  |
| Age (blood; years) | 71.5 (66.9-76.7) | 69.8 (65.8-75.3) | 73.4 (68.8-78.1) | 6.56E-11 |
| Sex |  |  |  | 0.508 |
| Female | 505 (50.6%) | 260 (51.6%) | 245 (49.5%) |  |
| Male | 494 (49.5%) | 244 (48.4%) | 250 (50.5%) |  |
| Education (years) | 16 (14-18) | 17 (16-18) | 16 (14-18) | 5.42E-04 |
| Race |  |  |  | 0.663 |
| White | 949 (95.5%) | 477 (95.0%) | 472 (95.9%) |  |
| B/AA | 39 (3.9%) | 21 (4.2%) | 18 (3.7%) |  |
| Asian | 6 (0.6%) | 4 (0.8%) | 2 (0.4%) |  |
| MMSE | 29 (27-30) | 29 (28-30) | 28 (25-29) | 4.64E-27 |
| Clinical diagnosis |  |  |  | 7.77E-27 |
| CU | 405 (40.5%) | 273 (54.2%) | 132 (26.7%) |  |
| MCI | 474 (47.5%) | 216 (42.9%) | 258 (52.1%) |  |
| Dementia | 120 (12.0%) | 15 (3.0%) | 105 (21.2%) |  |
| CDR-GS |  |  |  | 3.51E-21 |
| 0 | 405 (40.5%) | 272 (54.0%) | 133 (26.9%) |  |
| 0.5 | 527 (52.8%) | 224 (44.4%) | 303 (61.2%) |  |
| 1 | 66 (6.6%) | 8 (1.6%) | 58 (11.7%) |  |
| 2 | 1 (0.1%) | 0 | 1 (0.2%) |  |
| *APOE4* carrier |  |  |  | 3.18E-46 |
| No | 559 (56.0%) | 394 (78.2%) | 165 (33.3%) |  |
| Yes | 440 (44.0%) | 110 (21.8%) | 330 (66.7%) |  |
| Centiloid | 18 (0-74) | 0 (-7-7) | 74 (46-104) | 8.95E-165 |
| Tau PET |  |  |  | 4.51E-23 |
| Negative | 282 (79.4%) | 197 (98.0%) | 85 (55.2%) |  |
| Positive | 73 (20.6%) | 4 (2.0%) | 69 (44.8%) |  |
| Tau PET SUVR | 1.23 (1.17-1.34) | 1.20 (1.15-1.24) | 1.36 (1.24-1.63) | 7.50E-27 |

B/AA, Black/African-American; CU, cognitively unimpaired; MCI, mild cognitive impairment; PET, positron emission tomography; MMSE, Mini–Mental State Examination; CDR-GS, Clinical Dementia Rating global score; SUVR, standard uptake value ratio. Tau-PET SUVR is based on the meta-temporal region-of-interest SUVR.

**Table S6. Participant characteristics for the ADNI cohort stratified by tau-PET status.**

|  | **Total** | **Tau-PET-** | **Tau-PET+** | **p-value** |
| --- | --- | --- | --- | --- |
| Number of participants, n | 360 | 286 | 74 |  |
| Age (blood; years) | 70.0 (66.3-75.3) | 69.6 (66.3-75.3) | 71.3 (67.2-75.4) | 0.240 |
| Sex |  |  |  | 0.513 |
| Female | 197 (54.7%) | 159 (55.6%) | 38 (51.4%) |  |
| Male | 163 (45.3%) | 127 (44.4%) | 36 (48.7%) |  |
| Education (years) | 16 (15-18) | 16 (16-18) | 16 (14-18) | 0.025 |
| Race |  |  |  | 0.407 |
| White | 329 (92.7%) | 261 (92.6%) | 68 (93.2%) |  |
| B/AA | 20 (5.6%) | 15 (5.3%) | 5 (6.9%) |  |
| Asian | 6 (1.7%) | 6 (2.1%) | 0 |  |
| MMSE | 29 (27-30) | 29 (28-30) | 26 (23-28) | 7.21E-14 |
| Clinical diagnosis |  |  |  | 9.80E-19 |
| CU | 201 (55.8%) | 188 (65.7%) | 13 (17.6%) |  |
| MCI | 121 (33.6%) | 86 (30.1%) | 35 (47.3%) |  |
| Dementia | 38 (10.6%) | 12 (4.2%) | 26 (35.1%) |  |
| CDR-GS |  |  |  | 4.76E-15 |
| 0 | 202 (56.1%) | 189 (66.1%) | 13 (17.6%) |  |
| 0.5 | 138 (38.3%) | 90 (31.5%) | 48 (64.9%) |  |
| 1 | 20 (5.6%) | 7 (2.5%) | 13 (17.6%) |  |
| *APOE4* carrier |  |  |  | 5.90E-08 |
| No | 153 (42.5%) | 101 (35.3%) | 52 (70.3%) |  |
| Yes | 207 (57.5%) | 185 (64.7%) | 22 (29.7%) |  |
| Aβ PET |  |  |  | 4.51E-23 |
| Negative | 201 (56.6%) | 197 (69.9%) | 4 (5.5%) |  |
| Positive | 154 (43.4%) | 85 (30.1%) | 69 (94.5%) |  |
| Aβ-PET Centiloid | 11.0 (0-68.8) | 6 (-2-27) | 94 (74-118) | 3.76E-15 |
| Tau PET SUVR | 1.23 (1.17-1.34) | 1.21 (1.16-1.26) | 1.74 (1.47-2.16) | 2.47E-30 |

B/AA, Black/African-American; CU, cognitively unimpaired; MCI, mild cognitive impairment; PET, positron emission tomography; MMSE, Mini–Mental State Examination; CDR-GS, Clinical Dementia Rating global score; SUVR, standard uptake value ratio. Tau-PET SUVR is based on the meta-temporal region-of-interest SUVR.

**Table S7. Participant characteristics for the ADNI cohort stratified by clinical diagnosis.**

|  | **Total** | **CU** | **MCI** | **Dementia** | **p-value** |
| --- | --- | --- | --- | --- | --- |
| Number of participants, n | 1428 | 529 | 676 | 223 |  |
| Age (blood; years) | 72.8 (67.6-77.9) | 72.3 (67.8-77.0) | 72.4 (66.8-77.6) | 75.1 (69.7-80.2) | 7.89E-05 |
| Sex |  |  |  |  | 5.01E-10 |
| Female | 676 (47.3%) | 310 (58.6%) | 277 (41.0%) | 89 (39.9%) |  |
| Male | 752 (52.7%) | 219 (41.4%) | 399 (59.0%) | 134 (60.1%) |  |
| Education (years) | 16 (14-18) | 16 (15-18) | 16 (14-18) | 16 (14-18) | 4.13E-06 |
| Race |  |  |  |  | 0.022 |
| White | 1356 (95.3%) | 488 (92.8%) | 652 (96.7%) | 216 (96.9%) |  |
| B/AA | 56 (3.9%) | 33 (6.3%) | 16 (2.4%) | 7 (3.1%) |  |
| Asian | 11 (0.8%) | 5 (1.0%) | 6 (1.0%) | 0 |  |
| MMSE | 28 (26-30) | 29 (29-30) | 28 (26-29) | 23 (22-25) | 5.64E-142 |
| CDR-GS |  |  |  |  | 8.28E-194 |
| 0 | 526 (37.0%) | 522 (99.1%) | 4 (0.6%) | 0 |  |
| 0.5 | 784 (55.1%) | 5 (1.0%) | 669 (99.3%) | 110 (49.8%) |  |
| 1 | 111 (7.8%) | 0 | 1 (0.2%) | 110 (49.8%) |  |
| 2 | 1 (0.1%) | 0 | 0 | 1 (0.5%) |  |
| *APOE4* carrier |  |  |  |  | 5.77E-24 |
| No | 780 (54.6%) | 369 (69.8%) | 344 (50.9%) | 67 (30.0%) |  |
| Yes | 648 (45.4%) | 160 (30.3%) | 332 (49.1%) | 156 (70.0%) |  |
| Aβ PET |  |  |  |  | 7.77E-27 |
| Negative | 504 (50.5%) | 273 (67.4%) | 216 (45.6%) | 15 (12.5%) |  |
| Positive | 495 (49.6%) | 132 (32.6%) | 258 (54.4%) | 105 (87.5%) |  |
| Aβ-PET Centiloid | 5 (0-61) | 4 (-2-29) | 8 (0-66.5) | 4.5 (0-95.0) | 2.27E-08 |
| Tau PET |  |  |  |  | 9.80E-19 |
| Negative | 286 (79.4%) | 188 (93.5%) | 86 (71.1%) | 12 (21.6%) |  |
| Positive | 74 (20.6%) | 13 (6.5%) | 35 (28.9%) | 26 (68.4%) |  |
| Tau PET SUVR | 1.23 (1.17-1.34) | 1.21 (1.16-1.26) | 1.27 (1.18-1.42) | 1.66 (1.32-1.95) | 2.47E-23 |

B/AA, Black/African-American; CU, cognitively unimpaired; MCI, mild cognitive impairment; PET, positron emission tomography; MMSE, Mini–Mental State Examination; CDR-GS, Clinical Dementia Rating global score; SUVR, standard uptake value ratio. Tau-PET SUVR is based on the meta-temporal region-of-interest SUVR.

**Table S8. Baseline characteristics of cognitively unimpaired and mild cognitively impaired participants with longitudinal follow‑up in the ADRC cohort.**

|  | **Progression status for CU** | | | | **Progression status for MCI** | | | |
| --- | --- | --- | --- | --- | --- | --- | --- | --- |
|  | **Total** | **Stable** | **Progressor** | **p value** | **Total** | **Stable** | **Progressor** | **p value** |
| Number of participants, n | 310 | 250 | 60 |  | 230 | 79 | 151 |  |
| Age (baseline; years) | 69.5 (64.5-74.2) | 68.5 (64.2-73.5) | 72.6 (67.0-78.4) | 5.54E-04 | 73.3 (68.2-78.4) | 69.3 (62.3-75.6) | 74.4 (70.4-78.9) | 2.43E-06 |
| Years of follow-up | 8.8 (6.2-12.2) | 9.6 (7.1-13.2) | 5.2 (3.5-7.8) | 5.10E-15 |  |  |  |  |
| Sex |  |  |  | 8.81E-01 |  |  |  | 1.66E-01 |
| Female | 200 (64.5%) | 162 (64.8%) | 38 (63.3%) |  | 105 (45.7%) | 31 (39.2%) | 74 (49.0%) |  |
| Male | 110 (35.5%) | 88 (35.2%) | 22 (36.7%) |  | 125 (54.4%) | 48 (60.8%) | 77 (51.0%) |  |
| Education (years) | 16 (14-18) | 16 (14-18) | 14.5 (12-18) | 3.77E-03 | 16 (13-18) | 16 (14-18) | 16 (13-18) | 3.31E-01 |
| Race |  |  |  | 3.56E-02 |  |  |  | 7.18E-01 |
| NHW | 265 (85.5%) | 219 (87.6%) | 46 (76.7%) |  | 219 (95.2%) | 76 (96.2%) | 143 (94.7%) |  |
| B/AA | 43 (13.9%) | 29 (11.6%) | 14 (23.3%) |  | 8 (3.5%) | 2 (2.5%) | 6 (4.0%) |  |
| Others/missing | 2 (0.6%) | 2 (0.8%) |  |  | 3 (1.3%) | 1 (1.3%) | 2 (1.3%) |  |
| MMSE | 29 (28-30) | 29 (28-30) | 28 (26-29) | 2.08E-07 | 27 (25-28) | 28 (26.3-29) | 26 (24.3-27) | 9.23E-07 |
| CDR-GS (baseline) |  |  |  | 5.19E-05 |  |  |  |  |
| 0 | 237 (77.5%) | 203 (82.5%) | 34 (56.7%) |  | 7 (3.0%) | 4 (5.1%) | 3 (2.0%) | 2.67E-01 |
| 0.5 | 69 (22.6%) | 43 (17.5%) | 26 (43.3%) |  | 214 (93.0%) | 74 (93.7%) | 140 (92.7%) |  |
| 1 | 0 | 0 | 0 |  | 9 (3.9%) | 1 (1.3%) | 8 (5.3%) |  |
| Missing | 4 | 4 | 0 |  |  |  |  |  |
| *APOE4* carrier |  |  |  | 4.37E-02 |  |  |  | 3.64E-02 |
| No | 209 (67.6%) | 176 (70.4%) | 33 (55.9%) |  | 105 (45.9%) | 44 (55.7%) | 61 (40.7%) |  |
| Yes | 100 (32.4%) | 74 (29.6%) | 26 (44.1%) |  | 124 (54.2%) | 35 (44.3%) |  |  |
| Missing | 1 | 0 | 1 |  | 1 | 0 | 1 |  |

Abbreviation: NHW, Non-Hispanic White; B/AA, Black/African American; AD, Alzheimer’s disease; CU, clinically unimpaired; MCI, mild cognitive impairment; MMSE, Mini–Mental State Examination; CDR, Clinical Dementia Rating. Categorical variables are presented as count (percentage), and continuous variables as median and interquartile range. P‑values were calculated using the Kruskal–Wallis test for continuous variables and Fisher’s exact test or the chi‑square test for categorical variables. Note: Only participants who progressed within 5 years, or who had more than 5 years of follow‑up indicating stability within that period, were included in the progression‑status count.
